# Cross-Sectional Study on Philippine National Insurance Data on Malignancy

**DOI:** 10.1101/2022.11.17.22282434

**Authors:** S Alip, C Castillo, K. Ong, A J Castro, K Gonzales, G Gasa, P Fernandez, P Firaza, F Manalaysay, C Semblante, S Sergio, J Prodigalidad, M Macalalag, R Arcinas, A Roque

**Affiliations:** University of the Philippines, College of Medicine; Research Committee, Philippine Urological Association; Executive Council, Philippine Urological Association; Philippine Society of Urologic Oncologists; Philippine Endourological Society

## Abstract

**Introduction:** The purpose of the Urologic Diseases in the Philippines study is three-fold: to assess the burden of urologic disease in the country in prevalence and incidence, to identify orphan populations or underserved areas where urological care is most needed, and to provide an administrative data registry for which clinical data may be anchored on.

**Materials & Methods:** The data were then requested from the Corporate Planning Unit of PhilHealth, with the following inclusion parameters: all beneficiaries (members and their dependents) with a hospital claim in the years 2011 to 2021 with the following primary or secondary diagnoses e following variables: age, sex, admission date, outpatient/inpatient classification, specific illness code, claim amount, claim status, region and type of facility. Prevalence and incidence data were computed considering a 2-year lookback period. Health claims data is limited by limited clinical information it contains, and the selection bias of patients in frequent contact with the healthcare system.

**Results:** More than 30,000 health claims were reviewed. Incidence data for 2020, in comparison with the Globocan model is as follows: kidney cancer incidence (Philhealth 371 cases, Globocan model 2211) bladder cancer (Philhealth 261 cases, Globocan model 1,541), prostate cancer incidence (Philhealth 934, Globocan model 7,290), testicular cancer incidence (Philhealth 129, Globocan model 355), and penile cancer incidence (Philhealth 32, Globocan model 114). Other information such as prevalence data, regional and facility-type data are contained in the tables and supplementary material. Disparities in reporting may be due to remediable logistical and reporting issues, RVS and ICD exclusivity, and low service utilization.

**Conclusion:** Incidence across cancer types follow the trend of global estimates, with prostate cancer being the most prevalent, followed by kidney, bladder, testis, and penile cancers. For future studies, it is imperative that administrative and clinical data linkages are established to form a more holistic picture of the urologic disease burden in the country.

## Introduction

A broad tally of urologic diseases in the Philippines is necessary. As in most countries, available healthcare data is highly fragmented.^1^ Three major types of data sources are available to inform health research: clinical care data (including electronic health records, hospital or clinic-based datasets), large-scale administrative data (national insurance claims data, pharmaceutical data, government agency data), and clinical trial data (both publicly and privately funded). Access to administrative and clinical data, including linkages among them must be leveraged; opportunities to make data accessible at point of care for decision-making and health policy must be sought.

The purpose of the Urologic Diseases in the Philippines study is three-fold: to assess the burden of urologic disease in the country in prevalence and incidence, to identify orphan populations or underserved areas where urological care is most needed, and to provide an administrative data registry for which clinical data may be anchored on. The project is intended to be in several parts, each published separately as a rolling summary of the entirety of the project. The first part investigates the adequacy of a single national insurance claims database to inform urologic cancer epidemiology. The data from the Philippine Health Insurance Corporation (Philhealth) are meant to complement existing clinical data from cancer registries, and hospital data from the foremost urological training institutions.

There is no perfect database, and comprehensiveness is most often sacrificed for accessibility and ease of use. Currently, there are four subnational cancer registries in the Philippines that sample Manila, Rizal, Cebu, and Davao.^2^ Most Philippine data in global cancer catalogs, such as in the World Health Organization’s Global Cancer Registry, use statistical modeling for countries like the authors’ that lack a robust national registry, combining subnational data from the four registries and projections based on neighboring Southeast Asian countries.

Despite many arguable flaws and gaps in policy, it is high time that one of the strengths of the Universal Healthcare Act and national health insurance policy be utilized – that of universal enrolment and coverage, to gather a real-world estimate of healthcare utilization for cancer and other diseases. After the ratification of the Universal Healthcare Act (RA11223) in July of 2018, all Filipinos are now automatically included under the National Health Insurance Program, with coverage approaching 90% of the population.^3^

### Limitations of the Study

Using a single database of administrative data is fraught with inherent limitations. Firstly, there is an inherent bias in administrative data toward individuals who use health services. Secondly, the unit of analysis is a health claim (a clinic visit, admission, or contact with healthcare service), not individual patients. This data must be aggregated and disaggregated accordingly. Factors related to the natural history of the disease, such as disease recurrence, upstaging or morbidity, are impossible to generate. Thirdly, the usefulness of healthcare claims analyses can be complicated by incomplete health claims, unreliable or invalid coding, and missing information, such as important clinical variables. Diagnoses are often listed in the medical record; however, the principal diagnoses may not be cited accurately when the health claim is filed.

Specific limitations of this study include: a lack of direct supervision by primary or co-investigators of the Philhealth database: data are requested from the Philhealth representative in charge of searching and disaggregation and is forwarded to the investigators in a limited columnar file. Investigators did not have access to personal beneficiary data and no identifying patient information were released. Also, patient admissions recorded as ‘procedure only’ are captured by Relative Value Scale (RVS) code (not ICD code) and were not included in the study. Lookback capture was used to compensate for this underreporting.

Researchers dealing with a large volume of health claims information are always in an unsatisfactory position of having no definitive gold standard against which to validate administrative data.^4^ However, a prime advantage, of working with claims data is not only in quantifying the burden of disease, but in also defining a population for whom processes and outcomes of disease management may be explored. This is especially valuable in evaluating delivery and outcomes of care. Also, national databases mirror real-world data in that they reflect medical sociology and recording practices under economic constraints.^5^

As the recognized limitations of a claim or insurance-based databases are manifold, the authors’ fervent hope is to pave the way for more tailored, integrative, and accessible databases that may be relied upon by doctors and researchers alike.

## Materials and Methods

### Database Selection and Data Acquisition

In preparation for data collection and aggregation, a literature review of all existing studies on Philhealth-sourced data and concomitantly published datasets was done on Pubmed, HERDIN, and pre-print servers. No available datasets were identified. Local full texts articles, as well as Philhealth policy briefs and public releases were reviewed for information on unified Philhealth process flows. The research was registered under DOH-PHCRD Registration number PHRR220211-004331. The study procedures are in accordance with ICH-GCP principles, the provisions of the National Ethical Guidelines for Health and Health-related Research of 2017, and the Data Privacy Act of 2012 (RA 10173)

The data were then requested from the Corporate Planning Unit of PhilHealth, with the following inclusion parameters: all beneficiaries (members and their dependents) with a hospital claim in the years 2011 to 2021 with the following primary or secondary diagnoses: ICD-10 Codes (International Classification of Disease 10^th^ Edition) C60 malignant neoplasm of the penis, C61 malignant neoplasm of the prostate, C62 malignant neoplasm of the testis, C64 malignant neoplasm of the kidney, C67 malignant neoplasm of the bladder. ICD-10 coding was used as the 10^th^ edition coding is the current coding practice and has been for the inclusive sample years. Data retrieved from PhilHealth included the following variables: age, sex, admission date, outpatient/inpatient classification, specific illness code, claim amount, claim status. For patients with prostate cancer, data was further refined to include the following additional variables: region, and type of facility.

The patients are identified using a study-dedicated (anonymized) pseudo-identification number. Investigators had no access to identifying patient information. Each entry represented a single clinic visit. For instance, patients with multiple clinic visits were listed with the same pseudo-ID under multiple entries, with each entry representing each visit. Patients are unrestricted in their access to clinics, specialists, and hospitals. Thus, it is plausible for multiple claims to be made on the same day in a different clinic setting, or during multiple monthly visits to a single clinic. The first claim was chosen to define incidence. A disease-free interval of > 2 years was considered significant.

Denominators were taken from Philhealth annual reports and were verified through Philhealth correspondence. Philhealth releases public use data sets in one-year cycles. Gendered denominators for male genitourinary cancers were also supplied and refined by the Corporate Planning Unit.

### Statistical Analysis and the Lookback Period

Administrative claims data are heavily left and right censored. Proper selection of observable person time lends accuracy to estimates of prevalence and incidence, especially of chronic conditions. In this study, observable person-time was defined by enrollment in the claims database. Most contemporary studies on health claims utilize a lookback time to surveil the existing disease.^5–7^ Lookback periods should enable researchers to identify and exclude recurrent cases and increase the accuracy of the incidence estimation.^6^ If the condition of interest, in this study’s case cancer – a chronic condition, is observed to occur during the lookback time, then it is a prevalent disease. If the condition of interest is not observed to occur in the lookback time but does occur over the period of interest, then this case may be considered incident.^5^ The choice of lookback interval was critical to the analysis, and was defined as two years in this study, similar to previous studies on lookback for cancer.^8^

In this study, the denominator was the complete-period population or the number of beneficiaries (members and dependents eligible to claim) for the year of interest. Health claims data analysis involves calculation of diagnosis prevalence or incidence with the number of people with the primary diagnosis as the target illness, divided by the number of people covered by the health plan and not by those who have registered a health claim.^9^

All claims and health visits were aggregated under each unique patient identification number. That is to say, multiple visits of the same patient were aggregated under that patient and counted as one case under the first incident year. Frequency was used to described demographics of patients included in the study. Prevalence and incidence with their corresponding confidence intervals were calculated using the STATA software (command cii proportions) (StataCorp LLC, Texas, USA). Full operational definitions and formulae are available as supplementary material.

## Results

Table 1 shows the total registered Philhealth beneficiaries per year as a percentage of the Philippine National Statistics Authority population estimate. All Filipinos are automatically included under the National Health Insurance Program, registered beneficiaries are taken to mean qualified members based on entitlement. Beneficiaries include all registered members and dependents, regardless of premiums paid. It also shows total male beneficiaries, which served as total population-at-risk for prostate, testicular, and penile cancer. All membership categories are included in the count. Qualified dependents are declared by members and are similarly entitled to benefit availment.^3^

**Table 1.** Total Number of Registered Philhealth Beneficiaries

| Year | Total Registered Beneficiaries (Percentage of Population) |  | Total Male Beneficiaries |
| --- | --- | --- | --- |
| 2011 | 78,390,000 | (82.0%) | 14,702,032 (*39,222,568) |
| 2012 | 80,920,000. | (84.3%) | 15,834,192 (*41,040,780) |
| 2013 | 76,900,000. | (78.7%) | 16,945,352 (*38,984,123.7) |
| 2014 | 86,224,256. | (86.6%) | 44,403,501 |
| 2015 | 93,445,053. | (92.1%) | 47,814,214 |
| 2016 | 93,400,861 | (90.9%) | 47,960,271 |
| 2017 | 96,973,681 | (92.8%) | 46,307,729 |
| 2018 | 104,488,979 | (98.8%) | 51,486,361 |
| 2019 | 97,750,573. | (90.4%) | 46,681,095 |
| 2020 | 95,999,756 | (87.3%) | 46,024,677 |
| 2021 | 98,030,269. | (88.9%) | 45,746,463 |

\* based on the National Statistics Authority Population Estimate and Philhealth coverage for the year

The sudden increase in total male beneficiaries from 2013 to 2014 is due to the lack of sex-specific data in the earlier years for beneficiaries under categories including Citizen of Other Countries, Enterprise Owner, Family Driver, Filipino with Dual Citizenship, Government, Indigent, Informal Sector, *Kasambahay*, Naturalized Filipino Citizen, Organized Group, Sponsored. Although Philhealth started electronic claims processing in 1995, the electronic database was revamped in 2012 to accommodate the new payment mechanism.^10^ Subsequently, better data capture is apparent. Thus, for the years 2011-2013, the population-based estimate (with the respective coverage percentage applied) was used as the population-at-risk denominator instead (shown as a parenthetical in the corresponding rows).

The increase in coverage from 2017 to 2018 was due to the ratification of the Universal Health Care Act into law. Philippine national health insurance coverage is similar to, if not better than, neighboring countries of similar economic standing, such as Indonesia, which reported 75% coverage in 2018, and Vietnam, which reported 84% coverage in 2017^10^

### Kidney Cancer

A total of 3,729 unique patients were recorded in the Philhealth dataset where 392 patients had at least two different claim years (multiple patient visits). (**Figure K1**)

**Figure K1.**
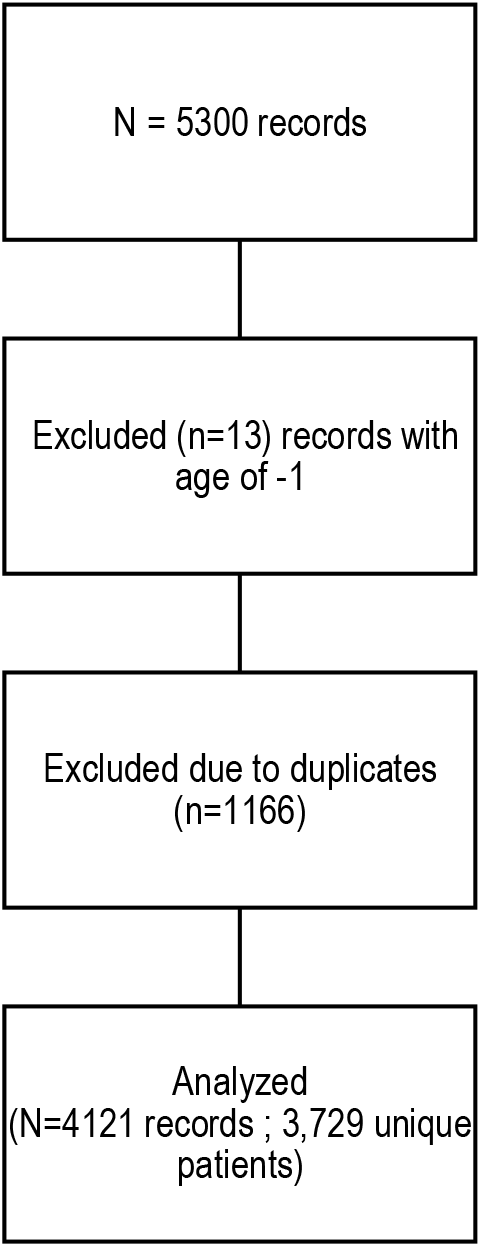
Distribution of kidney cancer patients from the 2011-2021 Philhealth database.

On the average, there were twice more male patients than females. Kidney cancer was most prevalent in 2019 followed by 2018 and 2017. The visits were also prevalent in the 40-59 age group and the patients were mostly males. (Table K1)

**Table K1.**
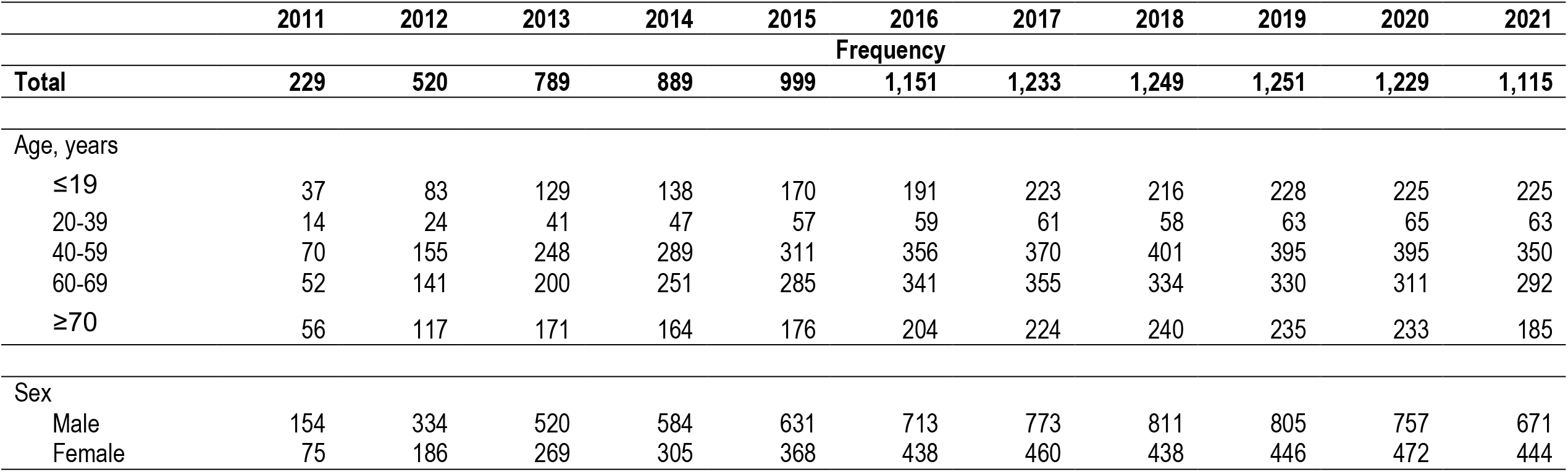
Prevalent Kidney Cancer Cases Per Year Based on Filed Claims Within Period of Interest and Lookback Period.

|  | 2011 | 2012 | 2013 | 2014 | 2015 | 2016 | 2017 | 2018 | 2019 | 2020 | 2021 |
| --- | --- | --- | --- | --- | --- | --- | --- | --- | --- | --- | --- |
|  | Frequency |  |  |  |  |  |  |  |  |  |  |
| **Total** | **229** | **520** | **789** | **889** | **999** | **1,151** | **1,233** | **1,249** | **1,251** | **1,229** | **1,115** |
| Age, years |  |  |  |  |  |  |  |  |  |  |  |
| ≤19 | 37 | 83 | 129 | 138 | 170 | 191 | 223 | 216 | 228 | 225 | 225 |
| 20-39 | 14 | 24 | 41 | 47 | 57 | 59 | 61 | 58 | 63 | 65 | 63 |
| 40-59 | 70 | 155 | 248 | 289 | 311 | 356 | 370 | 401 | 395 | 395 | 350 |
| 60-69 | 52 | 141 | 200 | 251 | 285 | 341 | 355 | 334 | 330 | 311 | 292 |
| ≥70 | 56 | 117 | 171 | 164 | 176 | 204 | 224 | 240 | 235 | 233 | 185 |
| Sex |  |  |  |  |  |  |  |  |  |  |  |
| Male | 154 | 334 | 520 | 584 | 631 | 713 | 773 | 811 | 805 | 757 | 671 |
| Female | 75 | 186 | 269 | 305 | 368 | 438 | 460 | 438 | 446 | 472 | 444 |

Excluded records with mistakes in recording (such as age, Figure K1) were usually part of denied claims.

Table K1 shows kidney cancer prevalence per year. As previously noted, prevalence is taken as the number of cases occurring in the year of interest, and the preceding two years (lookback period). Table K2 shows kidney cancer incidence per year. Incidence is taken as the number of new cases occurring in the year of interest. These cases are preceded by a disease-free period in the entirety of the two-year lookback period.

**Table K2.**
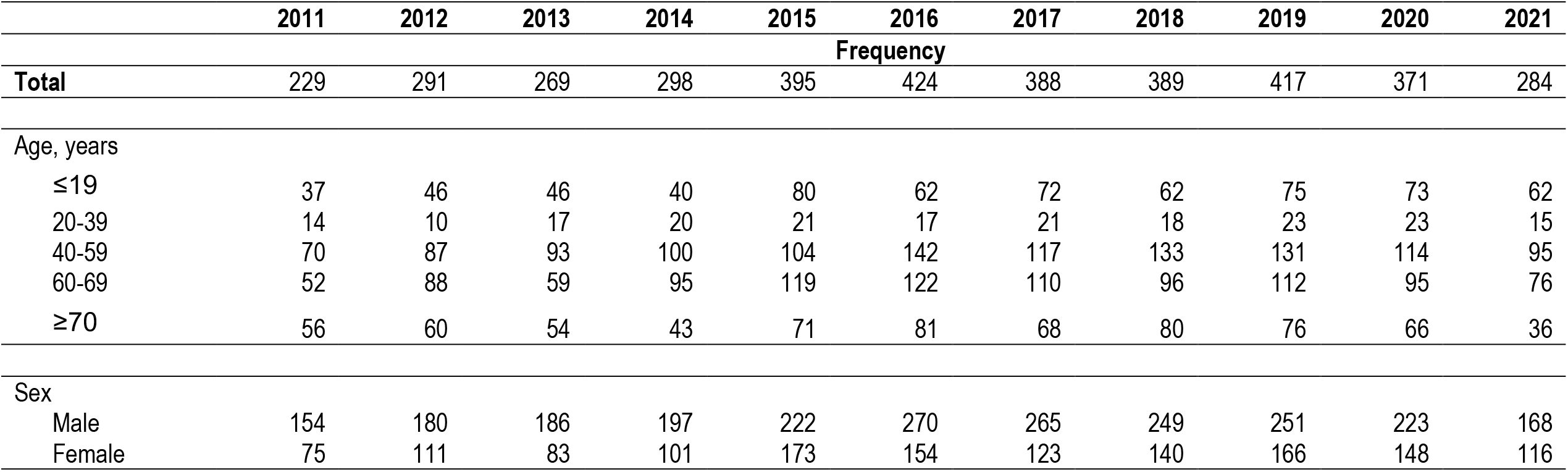
Incident Kidney Cancer Cases Per Year Based on Filed Claims Within Period of Interest.

Paid claims prevalence and incidence (denied and processing claims subtracted from total filed claims), and raw data without lookback periods are shown in Supplementary Tables 1 through 3. Incidence and prevalence rates across years are shown in Supplementary Table 4.

### Bladder Cancer

A total of 2,974 unique patients were recorded in the Philhealth dataset where 176 patients had at least two different claim years (multiple patient visits). (Figure B1)

**Figure B1.**
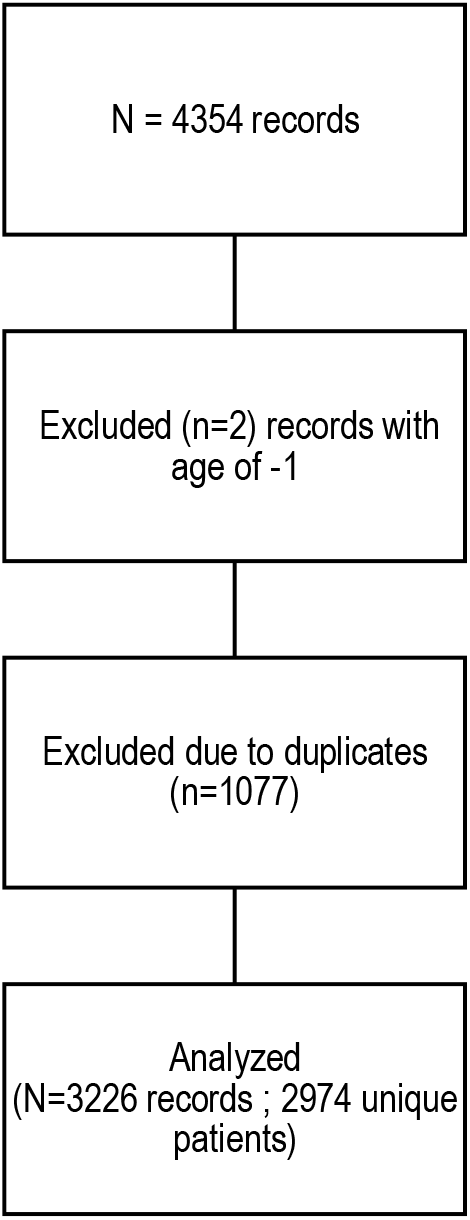
Distribution of bladder cancer patients from the 2011-2021 Philhealth database.

On the average, there were twice more male patients than females. Bladder cancer was most prevalent during 2017-2019. The visits were also prevalent in the older age group and the patients were mostly males. (Table B1) An unspecified bladder location was reported most often, followed by the trigone, and the internal urethra.

**Table B1.**
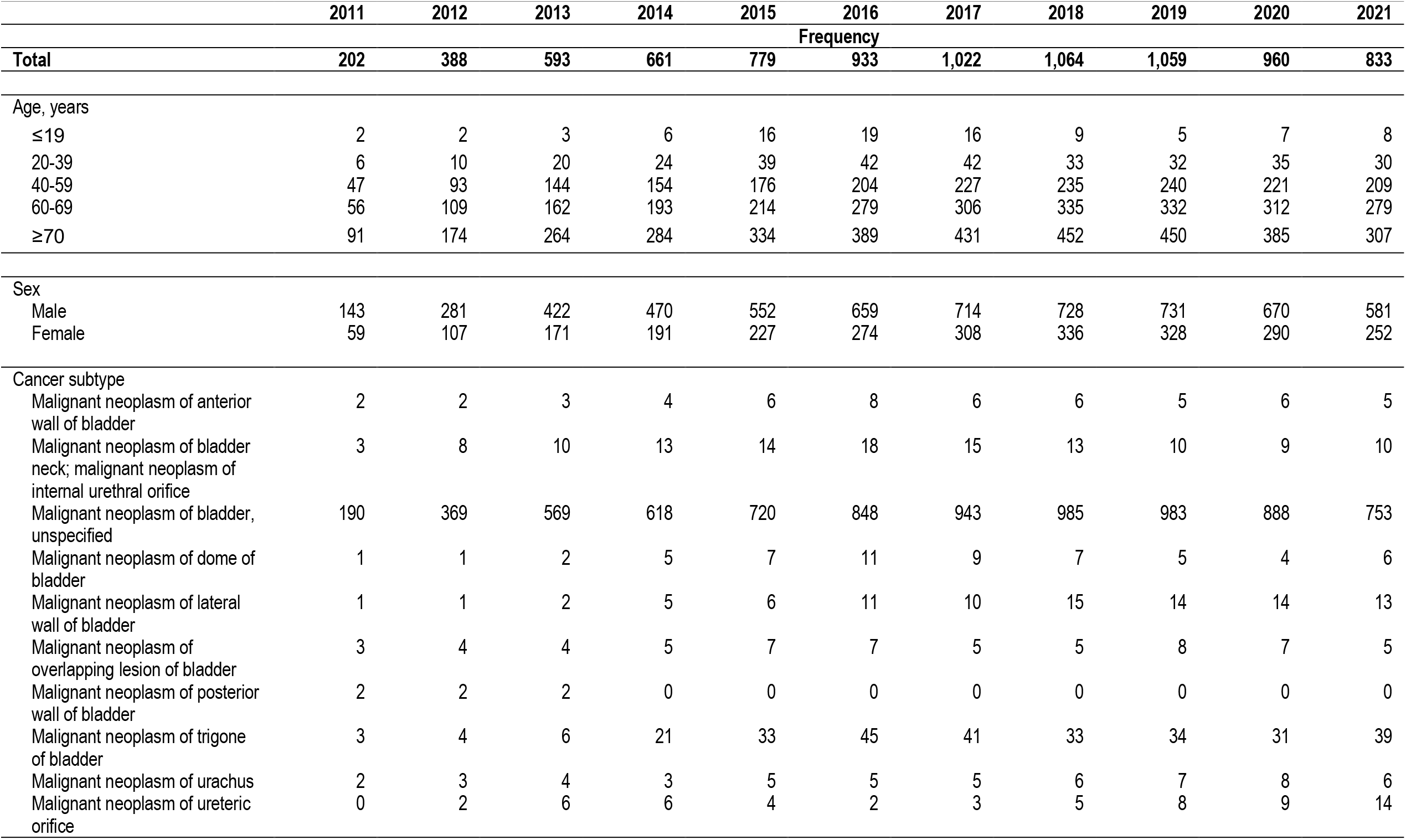
Prevalent Bladder Cancer Cases Per Year Based on Filed Claims Within Period of Interest and Lookback Period.

|  | 2011 | 2012 | 2013 | 2014 | 2015 | 2016 | 2017 | 2018 | 2019 | 2020 | 2021 |
| --- | --- | --- | --- | --- | --- | --- | --- | --- | --- | --- | --- |
|  | Frequency |  |  |  |  |  |  |  |  |  |  |
| **Total** | **202** | **388** | **593** | **661** | **779** | **933** | **1,022** | **1,064** | **1,059** | **960** | **833** |
| Age, years |  |  |  |  |  |  |  |  |  |  |  |
| ≤19 | 2 | 2 | 3 | 6 | 16 | 19 | 16 | 9 | 5 | 7 | 8 |
| 20-39 | 6 | 10 | 20 | 24 | 39 | 42 | 42 | 33 | 32 | 35 | 30 |
| 40-59 | 47 | 93 | 144 | 154 | 176 | 204 | 227 | 235 | 240 | 221 | 209 |
| 60-69 | 56 | 109 | 162 | 193 | 214 | 279 | 306 | 335 | 332 | 312 | 279 |
| ≥70 | 91 | 174 | 264 | 284 | 334 | 389 | 431 | 452 | 450 | 385 | 307 |
| Sex |  |  |  |  |  |  |  |  |  |  |  |
| Male | 143 | 281 | 422 | 470 | 552 | 659 | 714 | 728 | 731 | 670 | 581 |
| Female | 59 | 107 | 171 | 191 | 227 | 274 | 308 | 336 | 328 | 290 | 252 |
| Cancer subtype |  |  |  |  |  |  |  |  |  |  |  |
| Malignant neoplasm of anterior wall of bladder | 2 | 2 | 3 | 4 | 6 | 8 | 6 | 6 | 5 | 6 | 5 |
| Malignant neoplasm of bladder neck; malignant neoplasm of internal urethral orifice | 3 | 8 | 10 | 13 | 14 | 18 | 15 | 13 | 10 | 9 | 10 |
| Malignant neoplasm of bladder, unspecified | 190 | 369 | 569 | 618 | 720 | 848 | 943 | 985 | 983 | 888 | 753 |
| Malignant neoplasm of dome of bladder | 1 | 1 | 2 | 5 | 7 | 11 | 9 | 7 | 5 | 4 | 6 |
| Malignant neoplasm of lateral wall of bladder | 1 | 1 | 2 | 5 | 6 | 11 | 10 | 15 | 14 | 14 | 13 |
| Malignant neoplasm of overlapping lesion of bladder | 3 | 4 | 4 | 5 | 7 | 7 | 5 | 5 | 8 | 7 | 5 |
| Malignant neoplasm of posterior wall of bladder | 2 | 2 | 2 | 0 | 0 | 0 | 0 | 0 | 0 | 0 | 0 |
| Malignant neoplasm of trigone of bladder | 3 | 4 | 6 | 21 | 33 | 45 | 41 | 33 | 34 | 31 | 39 |
| Malignant neoplasm of urachus | 2 | 3 | 4 | 3 | 5 | 5 | 5 | 6 | 7 | 8 | 6 |
| Malignant neoplasm of ureteric orifice | 0 | 2 | 6 | 6 | 4 | 2 | 3 | 5 | 8 | 9 | 14 |

Excluded records with mistakes in recording (such as age, Figure 2) were usually part of denied claims.

Table B1 shows bladder cancer prevalence per year. As previously noted, prevalence is taken as the number of cases occurring in the year of interest and the preceding two years (lookback period). Table B2 shows bladder cancer incidence per year. Incidence is taken as the number of new cases occurring in the year of interest. These cases are preceded by a disease-free period in the entirety of the two-year lookback period.

**Table B2.** Incident Bladder Cancer Cases Per Year Based on Filed Claims Within Period of Interest.

|  | 2011 | 2012 | 2013 | 2014 | 2015 | 2016 | 2017 | 2018 | 2019 | 2020 | 2021 |
| --- | --- | --- | --- | --- | --- | --- | --- | --- | --- | --- | --- |
|  | Frequency |  |  |  |  |  |  |  |  |  |  |
| Total | 202 | 186 | 205 | 254 | 298 | 364 | 350 | 332 | 349 | 260 | 197 |
| Age, years |  |  |  |  |  |  |  |  |  |  |  |
| ≤19 | 2 | 0 | 1 | 4 | 11 | 4 | 1 | 2 | 1 | 4 | 3 |
| 20-39 | 6 | 4 | 10 | 10 | 18 | 13 | 10 | 10 | 12 | 10 | 7 |
| 40-59 | 47 | 46 | 51 | 50 | 68 | 80 | 79 | 70 | 83 | 63 | 58 |
| 60-69 | 56 | 53 | 53 | 83 | 73 | 118 | 111 | 104 | 115 | 89 | 69 |
| ≥70 | 91 | 83 | 90 | 107 | 128 | 149 | 149 | 146 | 138 | 94 | 60 |
| Sex |  |  |  |  |  |  |  |  |  |  |  |
| Male | 143 | 138 | 141 | 179 | 215 | 254 | 238 | 223 | 253 | 183 | 125 |
| Female | 59 | 48 | 64 | 75 | 83 | 110 | 112 | 109 | 96 | 77 | 72 |
| Cancer subtype |  |  |  |  |  |  |  |  |  |  |  |
| Malignant neoplasm of anterior wall of bladder | 2 | 0 | 1 | 3 | 2 | 3 | 1 | 2 | 2 | 2 | 1 |
| Malignant neoplasm of bladder neck; malignant neoplasm of internal urethral orifice | 3 | 5 | 2 | 6 | 6 | 6 | 3 | 4 | 3 | 2 | 5 |
| Malignant neoplasm of bladder, unspecified | 190 | 179 | 200 | 222 | 275 | 332 | 326 | 309 | 320 | 240 | 167 |
| Malignant neoplasm of dome of bladder | 1 | 0 | 1 | 4 | 2 | 5 | 2 | 0 | 3 | 1 | 2 |
| Malignant neoplasm of lateral wall of bladder | 1 | 0 | 1 | 4 | 1 | 6 | 3 | 6 | 5 | 3 | 5 |
| Malignant neoplasm of overlapping lesion of bladder | 3 | 1 | 0 | 4 | 3 | 0 | 2 | 3 | 3 | 1 | 1 |
| Malignant neoplasm of posterior wall of bladder | 2 | 0 | 0 | 0 | 0 | 0 | 0 | 0 | 0 | 0 | 0 |
| Malignant neoplasm of trigone of bladder | 3 | 1 | 2 | 18 | 13 | 14 | 14 | 5 | 15 | 11 | 13 |
| Malignant neoplasm of urachus | 2 | 1 | 1 | 1 | 2 | 2 | 1 | 3 | 3 | 2 | 0 |
| Malignant neoplasm of ureteric orifice | 0 | 2 | 4 | 0 | 0 | 2 | 1 | 2 | 5 | 2 | 7 |

Paid claims prevalence and incidence (denied and processing claims subtracted from total filed claims), and raw data without lookback periods are shown in Supplementary Tables 5 through 7. Incidence and prevalence rates across years are shown in Supplementary Table 8.

### Testicular Cancer

A total of 1,213 unique patients were recorded in the Philhealth dataset where 106 patients had at least two different claim years. Excluded records with mistakes in recording (such as age) were usually part of denied claims. (Figure T1)

**Figure T1.**
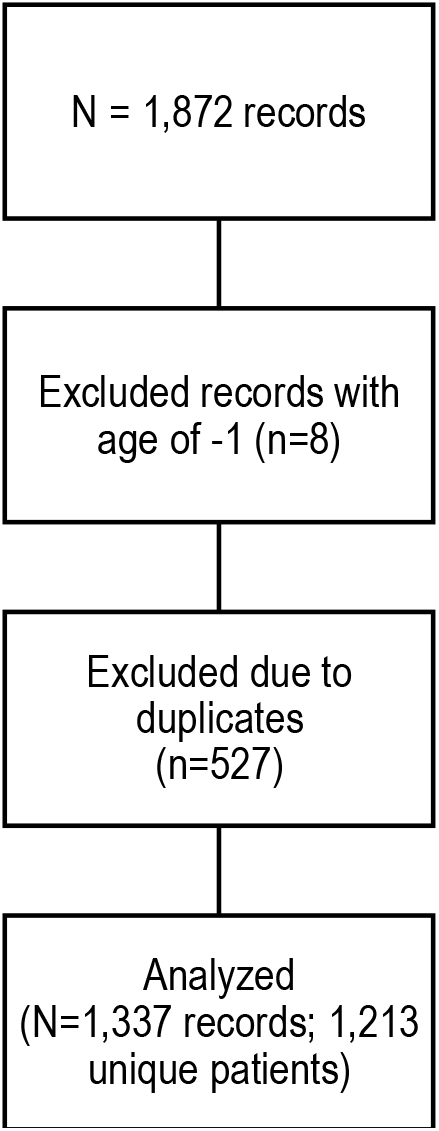
Distribution of testicular cancer patients from the 2011-2021 Philhealth database.

Considering a 2-year lookback, the years where testicular cancer has the most claims were on 2018 to 2020. (Table T1))

**Table T1.** Prevalent Testicular Cancer Cases Per Year Based on Filed Claims Within Period of Interest and Lookback Period.

|  | 2011 | 2012 | 2013 | 2014 | 2015 | 2016 | 2017 | 2018 | 2019 | 2020 | 2021 |
| --- | --- | --- | --- | --- | --- | --- | --- | --- | --- | --- | --- |
|  | Frequency |  |  |  |  |  |  |  |  |  |  |
| **Total** | **63** | **133** | **196** | **235** | **286** | **332** | **392** | **438** | **461** | **440** | **402** |
| Age, years |  |  |  |  |  |  |  |  |  |  |  |
| ≤19 | 15 | 32 | 44 | 61 | 74 | 97 | 119 | 124 | 135 | 131 | 126 |
| 20-39 | 31 | 60 | 85 | 90 | 113 | 136 | 157 | 188 | 185 | 185 | 172 |
| 40-59 | 14 | 35 | 53 | 62 | 70 | 69 | 81 | 80 | 93 | 85 | 82 |
| 60-69 | 3 | 5 | 10 | 13 | 18 | 19 | 19 | 25 | 24 | 23 | 13 |
| ≥70 | - | 1 | 4 | 9 | 11 | 11 | 16 | 21 | 24 | 16 | 9 |

Table T1 shows testicular cancer prevalence per year. As previously noted, prevalence is taken as the number of cases occurring in the year of interest, and the preceding two years (lookback period). Table T2 shows testicular cancer incidence per year. Incidence is taken as the number of new cases occurring in the year of interest. These cases are preceded by a disease-free period in the entirety of the two-year lookback period.

**Table T2.**
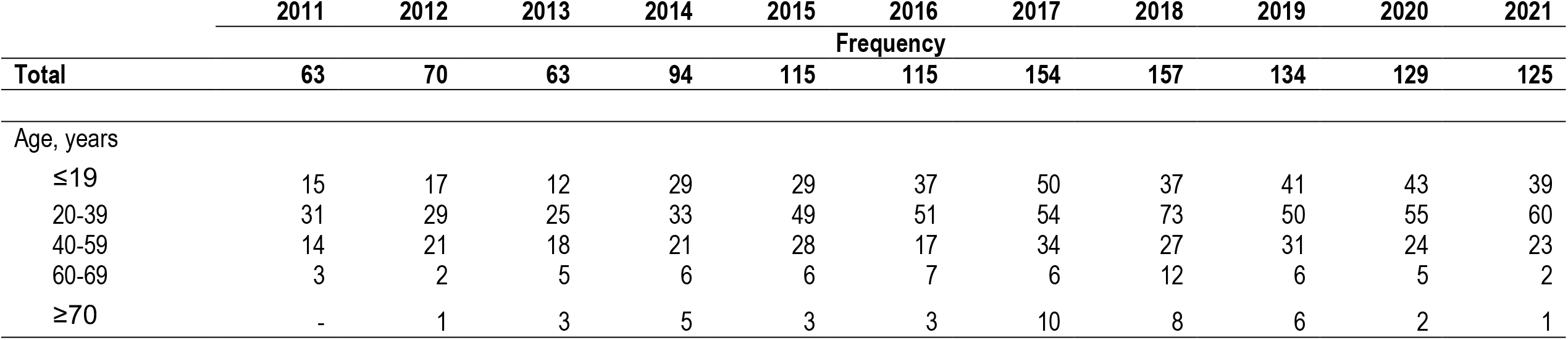
Incident Testicular Cancer Cases Per Year Based on Filed Claims Within Period of Interest.

Paid claims prevalence and incidence (denied and processing claims subtracted from total filed claims), and raw data without lookback periods are shown in Supplementary Tables 9 through 11. Incidence and prevalence rates across years are shown in Supplementary Table 12

### Penile Cancer

A total of 269 unique patients were recorded in the Philhealth dataset where 15 patients had at least two different claim years. (Figure PE1)

**Figure PE1.**
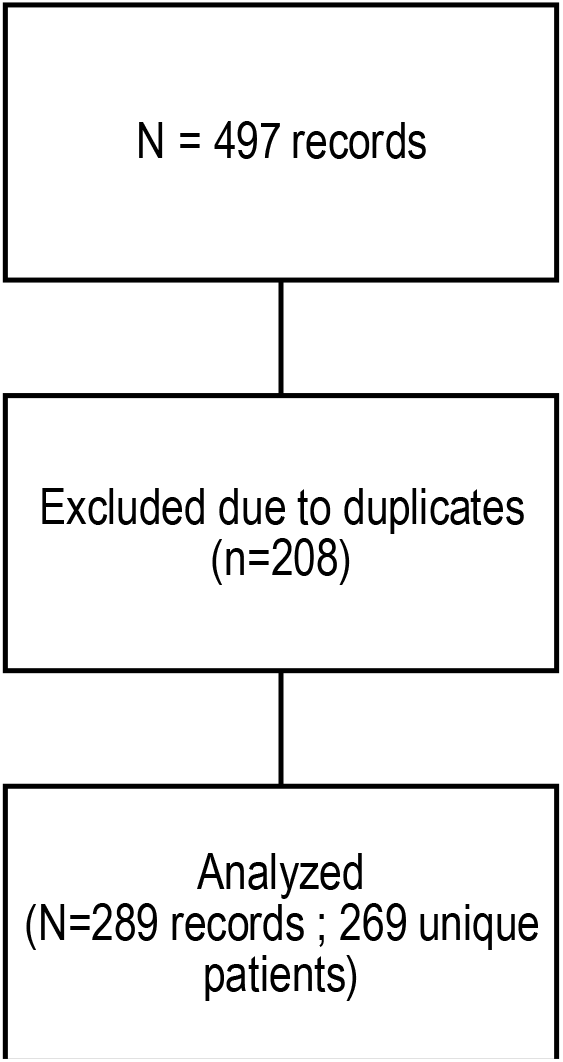
Distribution of penile cancer patients from the 2011-2021 Philhealth database

Considering a 2-year lookback, the year where penile cancer has the most claims was on 2020 followed by 2019 and 2017. (Table PE2)

Table PE1 shows penile cancer prevalence per year. As previously noted, prevalence is taken as the number of cases occurring in the year of interest, and the preceding two years (lookback period). Table PE2 shows penile cancer incidence per year. Incidence is taken as the number of new cases occurring in the year of interest. These cases are preceded by a disease-free period in the entirety of the two-year lookback period.

**Table PE1.** Prevalent Penile Cancer Cases Per Year Based on Filed Claims Within Period of Interest and Lookback Period.

|  | 2011 | 2012 | 2013 | 2014 | 2015 | 2016 | 2017 | 2018 | 2019 | 2020 | 2021 |
| --- | --- | --- | --- | --- | --- | --- | --- | --- | --- | --- | --- |
|  | Frequency |  |  |  |  |  |  |  |  |  |  |
| **Total** | **12** | **28** | **47** | **60** | **76** | **82** | **89** | **84** | **99** | **100** | **88** |
| Age, years |  |  |  |  |  |  |  |  |  |  |  |
| ≤19 | - | - | - | - | - | - | - | - | - | - | - |
| 20-39 | 3 | 3 | 6 | 8 | 11 | 10 | 11 | 10 | 14 | 15 | 14 |
| 40-59 | 5 | 13 | 25 | 34 | 43 | 43 | 46 | 47 | 52 | 55 | 46 |
| 60-69 | 1 | 5 | 7 | 9 | 13 | 14 | 18 | 14 | 20 | 17 | 17 |
| ≥70 | 3 | 7 | 9 | 9 | 9 | 15 | 14 | 13 | 13 | 13 | 11 |

**Table PE2.** Incident Penile Cancer Cases Per Year Based on Filed Claims Within Period of Interest.

|  | 2011 | 2012 | 2013 | 2014 | 2015 | 2016 | 2017 | 2018 | 2019 | 2020 | 2021 |
| --- | --- | --- | --- | --- | --- | --- | --- | --- | --- | --- | --- |
|  | Frequency |  |  |  |  |  |  |  |  |  |  |
| **Total** | **12** | **16** | **19** | **23** | **30** | **28** | **30** | **26** | **38** | **32** | **16** |
| Age, years |  |  |  |  |  |  |  |  |  |  |  |
| ≤19 | - | - | - | - | - | - | - | - | - | - | - |
| 20-39 | 3 | - | 3 | 4 | 4 | 2 | 5 | 3 | 5 | 7 | 2 |
| 40-59 | 5 | 8 | 12 | 13 | 16 | 13 | 16 | 18 | 17 | 20 | 9 |
| 60-69 | 1 | 4 | 2 | 3 | 6 | 5 | 7 | 2 | 10 | 3 | 4 |
| ≥70 | 3 | 4 | 2 | 3 | 4 | 8 | 2 | 3 | 6 | 2 | 1 |

Paid claims prevalence and incidence (denied and processing claims subtracted from total filed claims), and raw data without lookback periods are shown in Supplementary Tables 13 through 15. Incidence and prevalence rates across years are shown in Supplementary Table 16

### Prostate Cancer

A total of 10,910 unique patients were recorded in the Philhealth dataset where 1,289 patients had at least two different claim years. (Figure PR1)

**Figure PR1.**
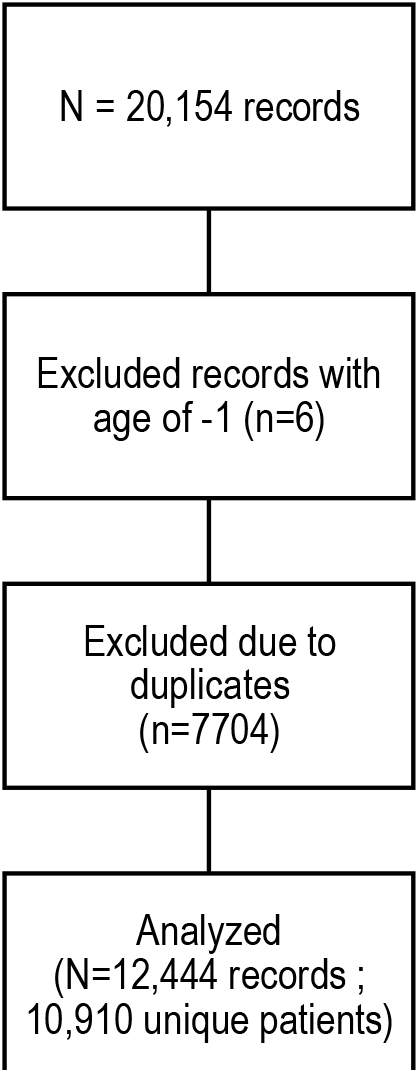
Distribution of prostate cancer patients from the 2011-2021 Philhealth database.

Considering a 2-year lookback, the years where prostate cancer has the most claims were on 2017 to 2019. (Table PR1) Most claims originate from NCR, followed by Central Luzon, Central Visayas, and Western Visayas. Most claims are from inpatients, seen at a tertiary or a secondary hospital.

**Table PR1.**
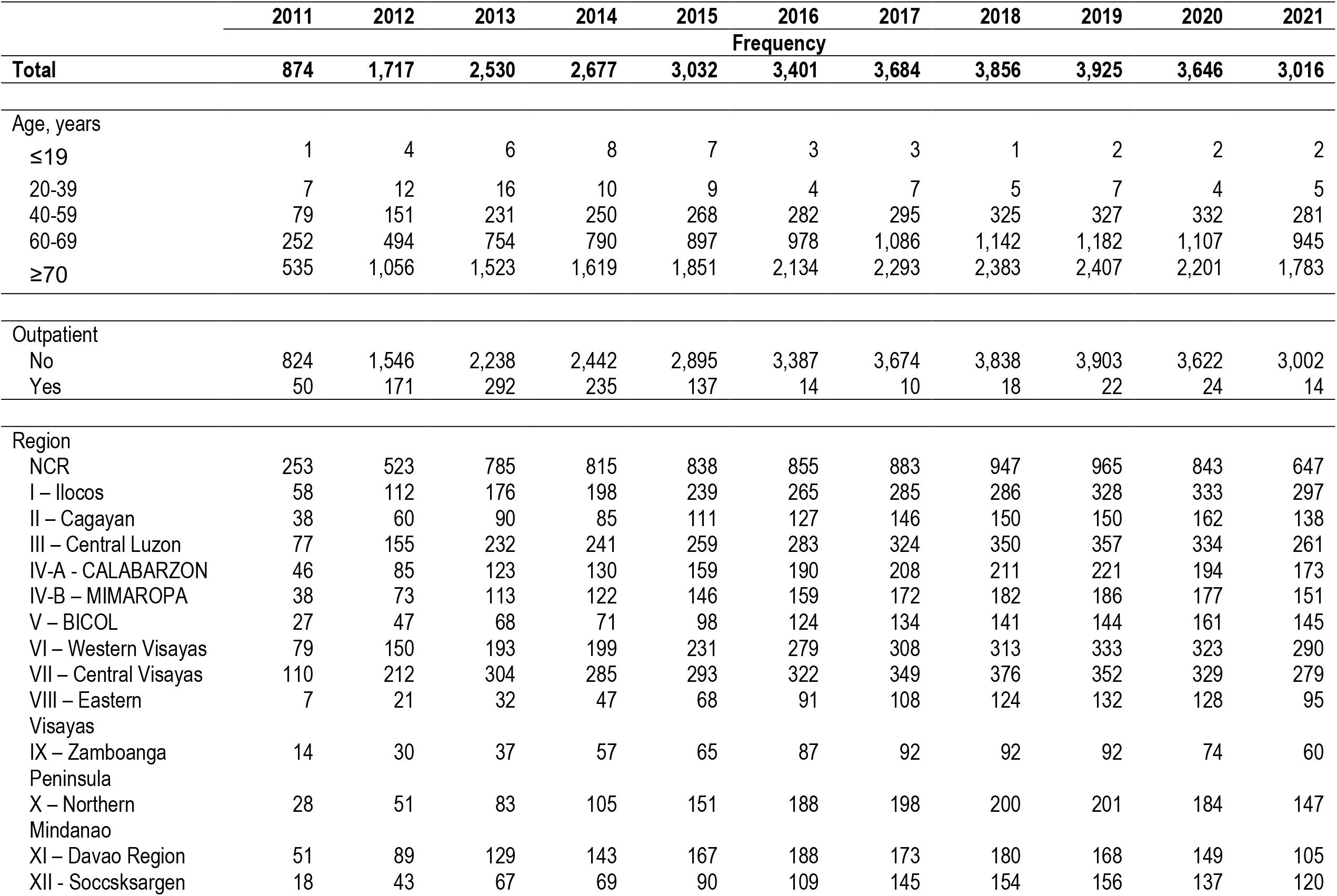
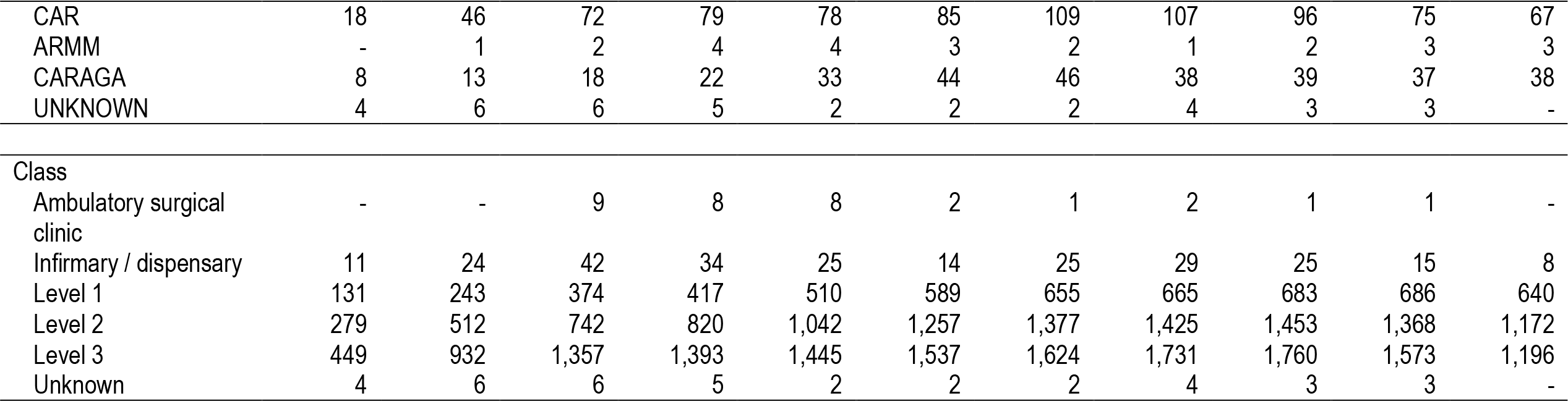
Prevalent Prostate Cancer Cases Per Year Based on Filed Claims Within Period of Interest and Lookback Period.

Table PR1 shows testicular cancer prevalence per year. As previously noted, prevalence is taken as the number of cases occurring in the year of interest, and the preceding two years (lookback period). Table PR2 shows kidney cancer incidence per year. Incidence is taken as the number of new cases occurring in the year of interest. These cases are preceded by a disease-free period in the entirety of the two-year lookback period.

**Table PR2.**
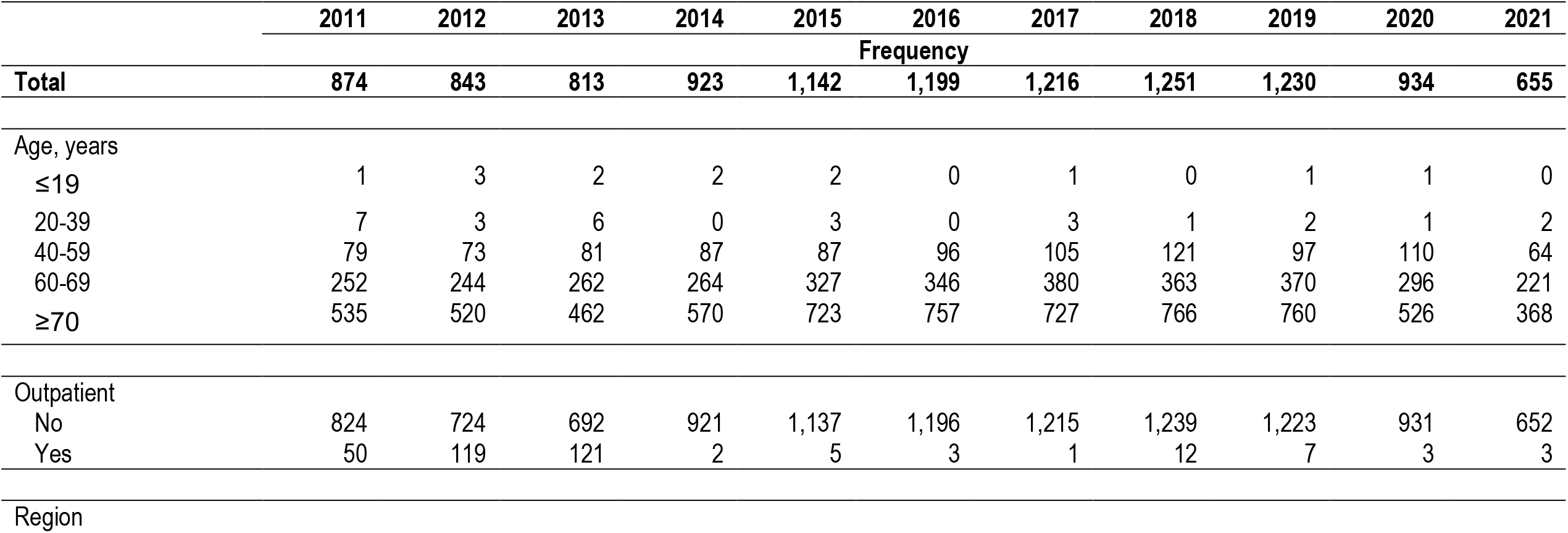
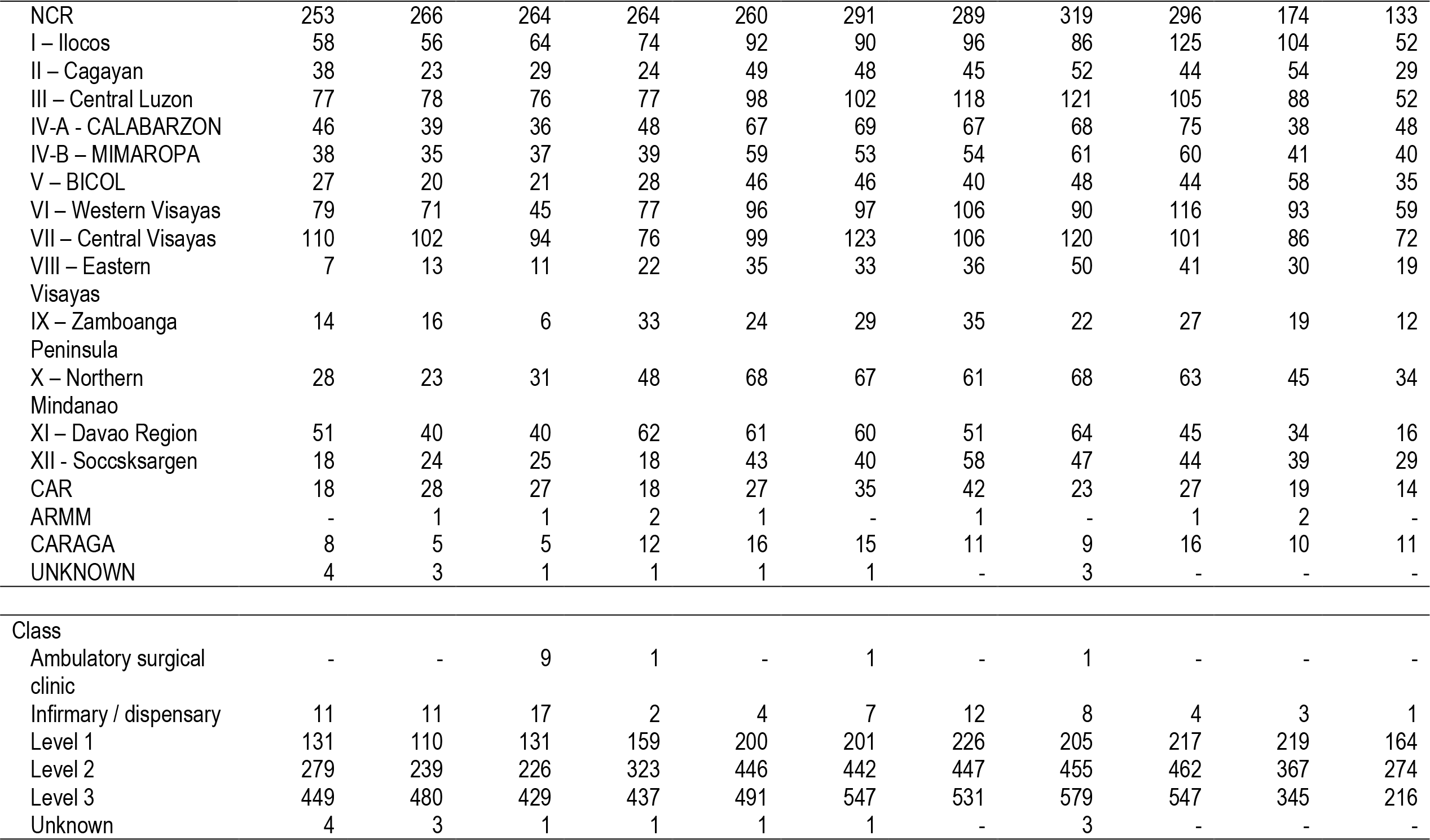
Incident Prostate Cancer Cases Per Year Based on Filed Claims Within Period of Interest.

|  | 2011 | 2012 | 2013 | 2014 | 2015 | 2016 | 2017 | 2018 | 2019 | 2020 | 2021 |
| --- | --- | --- | --- | --- | --- | --- | --- | --- | --- | --- | --- |
|  | Frequency |  |  |  |  |  |  |  |  |  |  |
| Total | 874 | 843 | 813 | 923 | 1,142 | 1,199 | 1,216 | 1,251 | 1,230 | 934 | 655 |
| Age, years |  |  |  |  |  |  |  |  |  |  |  |
| ≤19 | 1 | 3 | 2 | 2 | 2 | 0 | 1 | 0 | 1 | 1 | 0 |
| 20-39 | 7 | 3 | 6 | 0 | 3 | 0 | 3 | 1 | 2 | 1 | 2 |
| 40-59 | 79 | 73 | 81 | 87 | 87 | 96 | 105 | 121 | 97 | 110 | 64 |
| 60-69 | 252 | 244 | 262 | 264 | 327 | 346 | 380 | 363 | 370 | 296 | 221 |
| ≥70 | 535 | 520 | 462 | 570 | 723 | 757 | 727 | 766 | 760 | 526 | 368 |
| Outpatient |  |  |  |  |  |  |  |  |  |  |  |
| No | 824 | 724 | 692 | 921 | 1,137 | 1,196 | 1,215 | 1,239 | 1,223 | 931 | 652 |
| Yes | 50 | 119 | 121 | 2 | 5 | 3 | 1 | 12 | 7 | 3 | 3 |

|  | 2011 | 2012 | 2013 | 2014 | 2015 | 2016 | 2017 | 2018 | 2019 | 2020 | 2021 |
| --- | --- | --- | --- | --- | --- | --- | --- | --- | --- | --- | --- |
|  | Frequency |  |  |  |  |  |  |  |  |  |  |
| NCR | 253 | 266 | 264 | 264 | 260 | 291 | 289 | 319 | 296 | 174 | 133 |
| I – Ilocos | 58 | 56 | 64 | 74 | 92 | 90 | 96 | 86 | 125 | 104 | 52 |
| II – Cagayan | 38 | 23 | 29 | 24 | 49 | 48 | 45 | 52 | 44 | 54 | 29 |
| III – Central Luzon | 77 | 78 | 76 | 77 | 98 | 102 | 118 | 121 | 105 | 88 | 52 |
| IV-A - CALABARZON | 46 | 39 | 36 | 48 | 67 | 69 | 67 | 68 | 75 | 38 | 48 |
| IV-B – MIMAROPA | 38 | 35 | 37 | 39 | 59 | 53 | 54 | 61 | 60 | 41 | 40 |
| V – BICOL | 27 | 20 | 21 | 28 | 46 | 46 | 40 | 48 | 44 | 58 | 35 |
| VI – Western Visayas | 79 | 71 | 45 | 77 | 96 | 97 | 106 | 90 | 116 | 93 | 59 |
| VII – Central Visayas | 110 | 102 | 94 | 76 | 99 | 123 | 106 | 120 | 101 | 86 | 72 |
| VIII – Eastern Visayas | 7 | 13 | 11 | 22 | 35 | 33 | 36 | 50 | 41 | 30 | 19 |
| IX – Zamboanga Peninsula | 14 | 16 | 6 | 33 | 24 | 29 | 35 | 22 | 27 | 19 | 12 |
| X – Northern Mindanao | 28 | 23 | 31 | 48 | 68 | 67 | 61 | 68 | 63 | 45 | 34 |
| XI – Davao Region | 51 | 40 | 40 | 62 | 61 | 60 | 51 | 64 | 45 | 34 | 16 |
| XII - Soccsksargen | 18 | 24 | 25 | 18 | 43 | 40 | 58 | 47 | 44 | 39 | 29 |
| CAR | 18 | 28 | 27 | 18 | 27 | 35 | 42 | 23 | 27 | 19 | 14 |
| ARMM | - | 1 | 1 | 2 | 1 | - | 1 | - | 1 | 2 | - |
| CARAGA | 8 | 5 | 5 | 12 | 16 | 15 | 11 | 9 | 16 | 10 | 11 |
| UNKNOWN | 4 | 3 | 1 | 1 | 1 | 1 | - | 3 | - | - | - |
| **Class** |  |  |  |  |  |  |  |  |  |  |  |
| Ambulatory surgical clinic | - | - | 9 | 1 | - | 1 | - | 1 | - | - | - |
| Infirmery / dispensary | 11 | 11 | 17 | 2 | 4 | 7 | 12 | 8 | 4 | 3 | 1 |
| Level 1 | 131 | 110 | 131 | 159 | 200 | 201 | 226 | 205 | 217 | 219 | 164 |
| Level 2 | 279 | 239 | 226 | 323 | 446 | 442 | 447 | 455 | 462 | 367 | 274 |
| Level 3 | 449 | 480 | 429 | 437 | 491 | 547 | 531 | 579 | 547 | 345 | 216 |
| Unknown | 4 | 3 | 1 | 1 | 1 | 1 | - | 3 | - | - | - |

Paid claims prevalence and incidence (denied and processing claims subtracted from total filed claims), and raw data without lookback periods are shown in Supplementary Tables 17 through 19. Incidence and prevalence rates across years are shown in Supplementary Table 20

## Discussion

### Data Sources and Comparisons

At present there are four cancer registries in the Philippines, the Department of Health – Rizal Cancer Registry (DOH-RCR) with its catchment area totaling 1343 square kilometers; the Philippine Cancer Society – Manila Cancer Registry (PCS-MCR) with a total land catchment area of 266.5 square kilometers; the Cebu Cancer Registry with a catchment area of 793 square kilometers; and the Davao Cancer Registry with a catchment area estimate of 2,211.3 square kilometers.^11^ The former two registries have been the main source of cancer incidence data for more than three decades.^2^

The Global Cancer Observatory (Globocan), is a regular publication of worldwide cancer statistics by the International Agency for Research on Cancer (IARC), which is the specialized cancer agency of the World Health Organization. Globocan 2020 incidence figures for the Philippines extrapolate from the 2017 average of the three of the four abovementioned subnational registries (Cebu Cancer Registry, Manilla Cancer Registry, Rizal Cancer Registry) using prediction models, as historical Philippine cancer incidence data does not exist, and mortality data is not available.^12^

The Global Cancer Observatory publication for 2020 data includes the population-weighted average for incidence rates of the top five cancers in males and top five cancers in females in the Philippines. Prostate cancer in the third most incident cancer in males, the fifth most common overall for both sexes. For mortality data, cancer-specific data is not available, and incidence:mortality ratios are derived from cancer registry data in neighboring countries through modelling.^12^ Similarly, prevalence ratios use data from Nordic countries and are scaled using Human Development Index (HDI) ratios.

2020 Globocan Annual Incidence for prostate cancer is at 7,290, kidney at 2,211, bladder at 1,541, testis at 355, and penile at 114.^12^ Using cancer registry data, 2008 age-standardized national incidence of prostate cancer was 10.1 per 100,000 with the highest incidence rates in Quezon City, Paranaque, and Manila.^2^

On the other hand, Philhealth reports its coverage to be 90% of the entire population.^3^ Prior to 2018, Philhealth has publicly reported its coverage rate or entitlement rate based on premium payments. With the Universal Healthcare Act taking effect, current ‘registered members’ (the aforementioned 90%) are those already listed in the Philhealth database, having contact with the healthcare system, having secured their Member Data Record and Philhealth ID. To reiterate, unlike claims in Medicare or Medicaid of the United States which covers a certain group of individuals such as the elderly or the indigent population, the Philhealth insurance service has aimed to enroll 100% of the population. This representativeness and comprehensiveness afford us research opportunities not feasible using RCTs or single or oligo-institution databases (clinical data)^13^

Rather than viewing them as independent sources of data, administrative data and clinical registry data are often taken to complement each other to form a more holistic picture of the burden of illness. This has been done in various countries, such as the Medicare-SEER collaboration in the United States.^9^ Taking kidney cancer as an example, Philhealth data reports a 2020 incidence of 371 cases, while the Globocan estimate through registry extrapolation and modeling is pegged at 2211 cases.^12^ This five-fold difference reflects data loss from underreporting if the modeling estimates are taken as a gold standard. The underreporting may be due to remediable logistical issues: offline database status, deferral of Philhealth filing due to time or manpower constraints, neglect, and non-compliance. These errors are addressed by monitoring, incentives, and sanctions imposed by Philhealth, as part of institutional protocols.

The results expose one critical methodological limitation of this study. This is its failure to account for cases discharged as RVS (relative value scale) “procedure only” cases that are not co-listed under their respective ICD diagnosis. In its eschewing of the Fee for Service system, in favor of case rate payments in 2013, RVS and ICD classifications pathways are mutually exclusive, except for when claiming two case rates. The latter exceptions are subject to approval, and the more likely scenario is that claims are listed as either medical or procedural only.^14,15^ However, analysis is not as simple as merely including and adding these RVS cases. To illustrate, the RVS code for ‘cystoscopy, transurethral resection of bladder tumor’ may include benign cases. Conversely, RVS codes for ‘cystoscopy’ may include bladder cancer patients undergoing surveillance treatment. Clinical data – contained in dedicated registries – may be necessary to correctly classify cases. Future meticulously designed long-term studies are needed to address this data loss. Furthermore, standardization of treatment algorithms and respective reporting must be implemented, disseminated and adhered to by urologists.

Similar differences are seen across all cancers’ annual incidence: Bladder cancer (Philhealth 261 cases vs Globocan model 1,541), prostate cancer (Philhealth 934 vs Globocan model 7,290), testicular cancer (Philhealth 129 vs Globocan model 355), and penile cancer (Philhealth 32 vs Globocan model 114).

This study can also be viewed from the perspective of service utilization. In contrasting the Philippine estimate of cancer incidence in the global model to the number of those who enter the Philhealth system and avail themselves of healthcare services, the gap is largely apparent. Primarily due to factors already mentioned, but no doubt partly due to poor *de facto* access to government services in areas where awareness of Philhealth has yet to be reinforced.

## Conclusion

Claims analysis of urologic cancers may offer a reasonable estimate to guide service allocation. Incidence across cancer types follow the trend of global estimates, with prostate cancer being the most prevalent, followed by kidney, bladder, testis, and penile cancers. However, underreporting is inherent when the ICD inclusion criteria is applied alone. Low service utilization is evident, and urologists play an integral role in reinforcing national health insurance utilization. For future studies, it is imperative that administrative and clinical data linkages are established to form a more holistic picture of the urologic disease burden in the country. A rethinking of process flows for case reporting may also be considered to avoid data loss from patients who are coded as ‘procedure only’ cases; analyzing ICD and RVS codes hand-in-hand may serve to mitigate this loss.

## Data Availability

All data produced in the present study are available upon reasonable request to the authors

## Acknowledgments

The information herein was owed in part to the expert opinion of the following individuals and their constituent institutions

Philippine Endourological Society

Philippine Society of Urologic Oncology

Dr. Bernadette Lico and Mr. Paulus Bacud of the Philippine Health Insurance Corporation – Corporate Planning Unit

## A Appendix

### Supplementary Tables

**Supplementary Table 1.** Prevalent Kidney Cancer Cases Per Year Based on Paid Claims Only, Within Period of Interest and Lookback Period.

|  | 2011 | 2012 | 2013 | 2014 | 2015 | 2016 | 2017 | 2018 | 2019 | 2020 | 2021 |
| --- | --- | --- | --- | --- | --- | --- | --- | --- | --- | --- | --- |
|  | Frequency |  |  |  |  |  |  |  |  |  |  |
| Total | 223 | 507 | 769 | 870 | 977 | 1,130 | 1,201 | 1,208 | 1,183 | 1,154 | 980 |
| Age, years |  |  |  |  |  |  |  |  |  |  |  |
| ≤19 | 36 | 81 | 126 | 134 | 164 | 185 | 216 | 209 | 214 | 213 | 193 |
| 20-39 | 14 | 24 | 40 | 46 | 56 | 57 | 58 | 54 | 59 | 60 | 54 |
| 40-59 | 68 | 151 | 241 | 282 | 305 | 351 | 361 | 390 | 371 | 370 | 303 |
| 60-69 | 50 | 136 | 194 | 246 | 279 | 335 | 346 | 320 | 314 | 294 | 266 |
| ≥70 | 55 | 115 | 168 | 162 | 173 | 202 | 220 | 235 | 225 | 217 | 164 |
| Sex |  |  |  |  |  |  |  |  |  |  |  |
| Male | 149 | 323 | 504 | 568 | 615 | 698 | 754 | 783 | 763 | 709 | 600 |
| Female | 74 | 184 | 265 | 302 | 362 | 432 | 447 | 425 | 420 | 445 | 380 |

|  | 2011 | 2012 | 2013 | 2014 | 2015 | 2016 | 2017 | 2018 | 2019 | 2020 | 2021 |
| --- | --- | --- | --- | --- | --- | --- | --- | --- | --- | --- | --- |
|  | Frequency |  |  |  |  |  |  |  |  |  |  |
| **Total** | **223** | **507** | **769** | **870** | **977** | **1,130** | **1,201** | **1,208** | **1,183** | **1,154** | **980** |
| Age, years |  |  |  |  |  |  |  |  |  |  |  |
| ≤19 | 36 | 81 | 126 | 134 | 164 | 185 | 216 | 209 | 214 | 213 | 193 |
| 20-39 | 14 | 24 | 40 | 46 | 56 | 57 | 58 | 54 | 59 | 60 | 54 |
| 40-59 | 68 | 151 | 241 | 282 | 305 | 351 | 361 | 390 | 371 | 370 | 303 |
| 60-69 | 50 | 136 | 194 | 246 | 279 | 335 | 346 | 320 | 314 | 294 | 266 |
| ≥70 | 55 | 115 | 168 | 162 | 173 | 202 | 220 | 235 | 225 | 217 | 164 |
| Sex |  |  |  |  |  |  |  |  |  |  |  |
| Male | 149 | 323 | 504 | 568 | 615 | 698 | 754 | 783 | 763 | 709 | 600 |
| Female | 74 | 184 | 265 | 302 | 362 | 432 | 447 | 425 | 420 | 445 | 380 |

**Supplementary Table 2.**
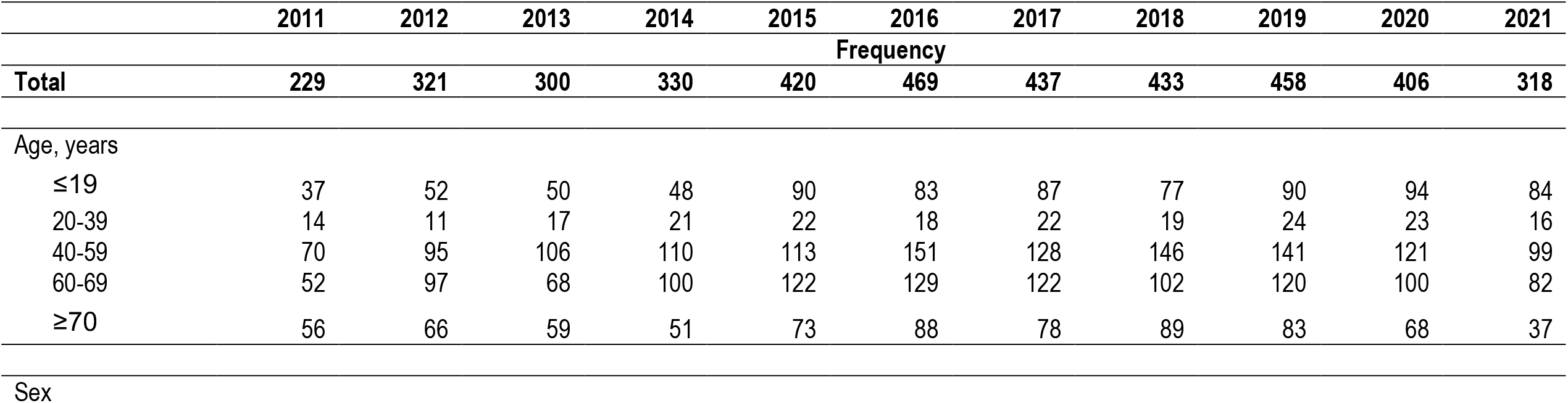
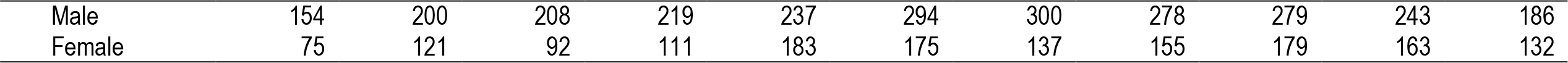
Prevalent Kidney Cancer Cases Per Year Based on Filed Claims Within Period of Interest Alone (Without Lookback)

**Supplementary Table 3.** Incident Kidney Cancer Cases Per Year Based on Paid Claims Only, Within Period of Interest.

|  | 2011 | 2012 | 2013 | 2014 | 2015 | 2016 | 2017 | 2018 | 2019 | 2020 | 2021 |
| --- | --- | --- | --- | --- | --- | --- | --- | --- | --- | --- | --- |
|  | Frequency |  |  |  |  |  |  |  |  |  |  |
| **Total** | **223** | **284** | **262** | **295** | **387** | **421** | **375** | **374** | **385** | **350** | **210** |
| Age, years |  |  |  |  |  |  |  |  |  |  |  |
| ≤19 | 36 | 45 | 45 | 38 | 77 | 64 | 69 | 62 | 67 | 72 | 43 |
| 20-39 | 14 | 10 | 16 | 20 | 21 | 15 | 20 | 17 | 21 | 21 | 10 |
| 40-59 | 68 | 85 | 90 | 100 | 104 | 140 | 112 | 130 | 117 | 109 | 69 |

|  | 2011 | 2012 | 2013 | 2014 | 2015 | 2016 | 2017 | 2018 | 2019 | 2020 | 2021 |
| --- | --- | --- | --- | --- | --- | --- | --- | --- | --- | --- | --- |
|  | Frequency |  |  |  |  |  |  |  |  |  |  |
| 60-69 | 50 | 85 | 58 | 94 | 115 | 121 | 108 | 88 | 108 | 90 | 60 |
| ≥70 | 55 | 59 | 53 | 43 | 70 | 81 | 66 | 77 | 72 | 58 | 28 |
| Sex |  |  |  |  |  |  |  |  |  |  |  |
| Male | 149 | 174 | 181 | 194 | 218 | 268 | 259 | 236 | 233 | 209 | 133 |
| Female | 74 | 110 | 81 | 101 | 169 | 153 | 116 | 138 | 152 | 141 | 77 |

**Supplementary Table 4.** Kidney Cancer Prevalence, Incidence Across Years.

| Year | Total beneficiaries | Prevalence of filed claims (95% CI) | Prevalence of paid claims (95% CI) | Incidence of filed claims (95% CI) | Incidence of paid claims (95% CI) |
| --- | --- | --- | --- | --- | --- |
| 2011 | 78,390,000 | 0.0000029 (0.0000026–0.0000033) | 0.0000028 (0.0000025–0.0000032) | 0.0000029 (0.0000026–0.0000033) | 0.0000028 (0.0000025–0.0000032) |
| 2012 | 80,920,000 | 0.0000064 (0.0000059–0.000007) | 0.0000063 (0.0000057–0.0000068) | 0.0000036 (0.0000032–0.000004) | 0.0000035 (0.0000031–0.0000039) |
| 2013 | 76,900,000 | 0.00001 (0.0000096–0.000011) | 0.00001 (0.0000093–0.000011) | 0.0000035 (0.0000031–0.0000039) | 0.0000034 (0.000003–0.0000039) |
| 2014 | 86,224,256 | 0.00001 (0.0000096–0.000011) | 0.00001 (0.0000094–0.000011) | 0.0000035 (0.0000031–0.0000039) | 0.0000034 (0.000003–0.0000038) |
| 2015 | 93,445,053 | 0.000011 (0.00001–0.000011) | 0.000011 (0.0000098–0.000011) | 0.0000042 (0.0000038–0.0000047) | 0.0000041 (0.0000037–0.0000046) |
| 2016 | 93,400,861 | 0.000012 (0.000012–0.000013) | 0.000012 (0.000011–0.000013) | 0.0000045 (0.0000041–0.000005) | 0.0000045 (0.0000041–0.000005) |
| 2017 | 96,973,681 | 0.000013 (0.000012–0.000013) | 0.000012 (0.000012–0.000013) | 0.000004 (0.0000036–0.0000044) | 0.0000039 (0.0000035–0.0000043) |
| 2018 | 104,488,979 | 0.000012 (0.000011–0.000013) | 0.000012 (0.000011–0.000012) | 0.0000037 (0.0000034–0.0000041) | 0.0000036 (0.0000032–0.000004) |
| 2019 | 97,750,573 | 0.000013 (0.000012–0.000014) | 0.000012 (0.000011–0.000013) | 0.0000043 (0.0000039–0.0000047) | 0.0000039 (0.0000036–0.0000044) |
| 2020 | 95,999,756 | 0.000013 (0.000012–0.000014) | 0.000012 (0.000011–0.000013) | 0.0000039 (0.0000035–0.0000043) | 0.0000037 (0.0000033–0.0000041) |
| 2021 | 98,030,269 | 0.000011 (0.000011–0.000012) | 0.00001 (0.0000094–0.000011) | 0.0000029 (0.0000026–0.0000033) | 0.0000021 (0.0000019–0.0000025) |

**Supplementary Table 5.** Prevalent Bladder Cancer Cases Per Year Based on Paid Claims Only Within Period of Interest and Lookback Period.

|  | 2011 | 2012 | 2013 | 2014 | 2015 | 2016 | 2017 | 2018 | 2019 | 2020 | 2021 |
| --- | --- | --- | --- | --- | --- | --- | --- | --- | --- | --- | --- |
|  | Frequency |  |  |  |  |  |  |  |  |  |  |
| Total | 191 | 364 | 562 | 637 | 760 | 913 | 992 | 1,026 | 1,006 | 893 | 733 |
| Age, years |  |  |  |  |  |  |  |  |  |  |  |
| ≤19 | 2 | 2 | 3 | 6 | 13 | 16 | 13 | 9 | 5 | 7 | 7 |
| 20-39 | 5 | 9 | 18 | 23 | 38 | 42 | 41 | 33 | 31 | 31 | 24 |
| 40-59 | 46 | 90 | 139 | 150 | 172 | 202 | 222 | 226 | 224 | 206 | 185 |
| 60-69 | 52 | 98 | 150 | 182 | 210 | 274 | 297 | 324 | 316 | 294 | 250 |

|  | 2011 | 2012 | 2013 | 2014 | 2015 | 2016 | 2017 | 2018 | 2019 | 2020 | 2021 |
| --- | --- | --- | --- | --- | --- | --- | --- | --- | --- | --- | --- |
|  | Frequency |  |  |  |  |  |  |  |  |  |  |
| ≥70 | 86 | 165 | 252 | 276 | 327 | 379 | 419 | 434 | 430 | 355 | 267 |
| Sex |  |  |  |  |  |  |  |  |  |  |  |
| Male | 137 | 264 | 399 | 449 | 538 | 647 | 695 | 701 | 691 | 623 | 518 |
| Female | 54 | 100 | 163 | 188 | 222 | 266 | 297 | 325 | 315 | 270 | 215 |
| Cancer subtype (not mutually exclusive) |  |  |  |  |  |  |  |  |  |  |  |
| Malignant neoplasm of anterior wall of bladder | 2 | 0 | 1 | 3 | 2 | 3 | 1 | 2 | 2 | 1 | 1 |
| Malignant neoplasm of bladder neck; malignant neoplasm of internal urethral orifice | 3 | 5 | 2 | 6 | 6 | 4 | 3 | 4 | 3 | 2 | 5 |
| Malignant neoplasm of bladder, unspecified | 180 | 181 | 215 | 236 | 274 | 334 | 342 | 302 | 318 | 237 | 138 |
| Malignant neoplasm of dome of bladder | 1 | 0 | 1 | 3 | 2 | 5 | 2 | 0 | 3 | 1 | 2 |
| Malignant neoplasm of lateral wall of bladder | 1 | 0 | 1 | 4 | 1 | 6 | 3 | 6 | 4 | 3 | 3 |
| Malignant neoplasm of overlapping lesion of bladder | 3 | 1 | 0 | 4 | 2 | 0 | 2 | 3 | 3 | 1 | 1 |
| Malignant neoplasm of posterior wall of bladder | 1 | 0 | 0 | 0 | 0 | 0 | 0 | 0 | 0 | 0 | 0 |
| Malignant neoplasm of trigone of bladder | 3 | 1 | 1 | 18 | 13 | 14 | 14 | 5 | 13 | 9 | 7 |
| Malignant neoplasm of urachus | 2 | 1 | 2 | 1 | 2 | 2 | 1 | 3 | 3 | 3 | 1 |
| Malignant neoplasm of ureteric orifice | 0 | 2 | 4 | 0 | 0 | 2 | 1 | 2 | 5 | 2 | 3 |

**Supplementary Table 6.** Prevalent Bladder Cancer Cases Per Year Based on Filed Claims Within Period of Interest Along (Without Lookback Period)

|  | 2011 | 2012 | 2013 | 2014 | 2015 | 2016 | 2017 | 2018 | 2019 | 2020 | 2021 |
| --- | --- | --- | --- | --- | --- | --- | --- | --- | --- | --- | --- |
|  | Frequency |  |  |  |  |  |  |  |  |  |  |
| Total | 202 | 201 | 225 | 270 | 308 | 373 | 378 | 345 | 371 | 280 | 209 |

|  | 2011 | 2012 | 2013 | 2014 | 2015 | 2016 | 2017 | 2018 | 2019 | 2020 | 2021 |
| --- | --- | --- | --- | --- | --- | --- | --- | --- | --- | --- | --- |
|  | Frequency |  |  |  |  |  |  |  |  |  |  |
| ≤19 | 2 | 1 | 1 | 4 | 11 | 6 | 2 | 2 | 1 | 4 | 4 |
| 20-39 | 6 | 4 | 11 | 11 | 19 | 13 | 10 | 12 | 13 | 10 | 7 |
| 40-59 | 47 | 53 | 58 | 56 | 68 | 84 | 87 | 75 | 87 | 67 | 61 |
| 60-69 | 56 | 57 | 58 | 87 | 79 | 120 | 114 | 109 | 120 | 96 | 72 |
| ≥70 | 91 | 86 | 97 | 112 | 131 | 150 | 165 | 147 | 150 | 103 | 65 |
| Sex |  |  |  |  |  |  |  |  |  |  |  |
| Male | 143 | 150 | 157 | 189 | 222 | 262 | 255 | 230 | 270 | 196 | 133 |
| Female | 59 | 51 | 68 | 81 | 86 | 111 | 123 | 115 | 101 | 84 | 76 |
| Cancer subtype |  |  |  |  |  |  |  |  |  |  |  |
| Malignant neoplasm of anterior wall of bladder | 2 | 0 | 1 | 3 | 2 | 3 | 1 | 2 | 2 | 2 | 1 |
| Malignant neoplasm of bladder neck; malignant neoplasm of internal urethral orifice | 3 | 5 | 2 | 6 | 6 | 6 | 3 | 4 | 3 | 2 | 5 |
| Malignant neoplasm of bladder, unspecified | 190 | 194 | 221 | 240 | 285 | 341 | 354 | 322 | 342 | 258 | 179 |
| Malignant neoplasm of dome of bladder | 1 | 0 | 1 | 4 | 2 | 5 | 2 | 0 | 3 | 1 | 2 |
| Malignant neoplasm of lateral wall of bladder | 1 | 0 | 1 | 4 | 1 | 6 | 3 | 6 | 5 | 3 | 5 |
| Malignant neoplasm of overlapping lesion of bladder | 3 | 1 | 0 | 4 | 3 | 0 | 2 | 3 | 3 | 1 | 1 |
| Malignant neoplasm of posterior wall of bladder | 2 | 0 | 0 | 0 | 0 | 0 | 0 | 0 | 0 | 0 | 0 |
| Malignant neoplasm of trigone of bladder | 3 | 1 | 2 | 18 | 13 | 14 | 14 | 5 | 15 | 12 | 13 |
| Malignant neoplasm of urachus | 2 | 1 | 2 | 1 | 2 | 2 | 1 | 3 | 3 | 4 | 1 |
| Malignant neoplasm of ureteric orifice | 0 | 2 | 4 | 0 | 0 | 2 | 1 | 2 | 5 | 2 | 7 |

**Supplementary Table 7.** Incident Bladder Cancer Cases Per Year Based on Paid Claims Only, Within Period of Interest and Lookback Period.

|  | 2011 | 2012 | 2013 | 2014 | 2015 | 2016 | 2017 | 2018 | 2019 | 2020 | 2021 |
| --- | --- | --- | --- | --- | --- | --- | --- | --- | --- | --- | --- |
|  | Frequency |  |  |  |  |  |  |  |  |  |  |
| **Total** | **196** | **176** | **205** | **257** | **295** | **362** | **343** | **318** | **335** | **242** | **149** |
| Age, years |  |  |  |  |  |  |  |  |  |  |  |
| ≤19 | 2 | 0 | 1 | 4 | 8 | 4 | 1 | 2 | 1 | 4 | 2 |
| 20-39 | 5 | 4 | 9 | 10 | 18 | 13 | 10 | 10 | 11 | 7 | 5 |
| 40-59 | 46 | 44 | 49 | 50 | 66 | 80 | 76 | 66 | 75 | 61 | 44 |
| 60-69 | 52 | 46 | 52 | 80 | 73 | 116 | 107 | 100 | 108 | 84 | 53 |
| ≥70 | 86 | 79 | 87 | 106 | 125 | 143 | 146 | 138 | 130 | 82 | 42 |
| Sex |  |  |  |  |  |  |  |  |  |  |  |
| Male | 137 | 127 | 135 | 175 | 211 | 250 | 230 | 211 | 235 | 168 | 97 |
| Female | 54 | 46 | 63 | 75 | 79 | 106 | 110 | 105 | 90 | 70 | 49 |
| Cancer subtype |  |  |  |  |  |  |  |  |  |  |  |
| Malignant neoplasm of anterior wall of bladder | 2 | 0 | 1 | 3 | 2 | 3 | 1 | 2 | 2 | 1 | 1 |
| Malignant neoplasm of bladder neck; malignant neoplasm of internal urethral orifice | 3 | 5 | 2 | 6 | 6 | 4 | 3 | 4 | 3 | 2 | 5 |
| Malignant neoplasm of bladder, unspecified | 180 | 166 | 194 | 218 | 267 | 326 | 316 | 293 | 299 | 223 | 127 |
| Malignant neoplasm of dome of bladder | 1 | 0 | 1 | 3 | 2 | 5 | 2 | 0 | 3 | 1 | 2 |
| Malignant neoplasm of lateral wall of bladder | 1 | 0 | 1 | 4 | 1 | 6 | 3 | 6 | 4 | 3 | 3 |
| Malignant neoplasm of overlapping lesion of bladder | 3 | 1 | 0 | 4 | 2 | 0 | 2 | 3 | 3 | 1 | 1 |
| Malignant neoplasm of posterior wall of bladder | 1 | 0 | 0 | 0 | 0 | 0 | 0 | 0 | 0 | 0 | 0 |
| Malignant neoplasm of trigone of bladder | 3 | 1 | 1 | 18 | 13 | 14 | 14 | 5 | 13 | 8 | 7 |
| Malignant neoplasm of urachus | 2 | 1 | 1 | 1 | 2 | 2 | 1 | 3 | 3 | 1 | 0 |
| Malignant neoplasm of ureteric orifice | 0 | 2 | 4 | 0 | 0 | 2 | 1 | 2 | 5 | 2 | 3 |

**Supplementary Table 8.** Bladder Cancer Prevalence, Incidence Across Years.

| Year | Total beneficiaries | Prevalence of filed claims (95% CI) | Prevalence of paid claims (95% CI) | Incidence of filed claims (95% CI) | Incidence of paid claims (95% CI) |
| --- | --- | --- | --- | --- | --- |
| 2011 | 78,390,000 | 0.0000026 (0.0000022–0.000003) | 0.0000024 (0.0000021–0.0000028) | 0.0000026 (0.0000022–0.000003) | 0.0000024 (0.0000021–0.0000028) |
| 2012 | 80,920,000 | 0.0000048 (0.0000043–0.0000053) | 0.0000045 (0.0000041–0.000005) | 0.0000023 (0.000002–0.0000027) | 0.0000021 (0.0000018–0.0000025) |
| 2013 | 76,900,000 | 0.0000077 (0.0000071–0.0000084) | 0.0000073 (0.0000067–0.0000079) | 0.0000027 (0.0000023–0.0000031) | 0.0000026 (0.0000022–0.000003) |
| 2014 | 86,224,256 | 0.0000077 (0.0000071–0.0000083) | 0.0000074 (0.0000068–0.000008) | 0.000003 (0.0000026–0.0000033) | 0.0000029 (0.0000026–0.0000033) |
| 2015 | 93,445,053 | 0.0000083 (0.0000078–0.0000089) | 0.0000081 (0.0000076–0.0000087) | 0.0000032 (0.0000028–0.0000036) | 0.0000031 (0.0000028–0.0000035) |
| 2016 | 93,400,861 | 0.00001 (0.0000094–0.000011) | 0.0000098 (0.0000092–0.00001) | 0.0000039 (0.0000035–0.0000043) | 0.0000038 (0.0000034–0.0000042) |
| 2017 | 96,973,681 | 0.000011 (0.0000099–0.000011) | 0.00001 (0.0000096–0.000011) | 0.0000036 (0.0000032–0.000004) | 0.0000035 (0.0000031–0.0000039) |
| 2018 | 104,488,979 | 0.00001 (0.0000096–0.000011) | 0.0000098 (0.0000092–0.00001) | 0.0000032 (0.0000028–0.0000035) | 0.000003 (0.0000027–0.0000034) |
| 2019 | 97,750,573 | 0.000011 (0.00001–0.000012) | 0.00001 (0.0000097–0.000011) | 0.0000036 (0.0000032–0.000004) | 0.0000033 (0.000003–0.0000037) |
| 2020 | 95,999,756 | 0.00001 (0.0000094–0.000011) | 0.0000093 (0.0000087–0.0000099) | 0.0000027 (0.0000024–0.0000031) | 0.0000025 (0.0000022–0.0000028) |
| 2021 | 98,030,269 | 0.0000085 (0.0000079–0.0000091) | 0.0000075 (0.000007–0.000008) | 0.000002 (0.0000017–0.0000023) | 0.0000015 (0.0000013–0.0000018) |

**Supplementary Table 9.** Prevalent Testicular Cancer Cases Per Year Based on Paid Claims Only, Within Period of Interest and Lookback Period.

|  | 2011 | 2012 | 2013 | 2014 | 2015 | 2016 | 2017 | 2018 | 2019 | 2020 | 2021 |
| --- | --- | --- | --- | --- | --- | --- | --- | --- | --- | --- | --- |
|  | Frequency |  |  |  |  |  |  |  |  |  |  |
| Total | 61 | 130 | 192 | 231 | 281 | 328 | 387 | 425 | 432 | 409 | 358 |
| Age, years |  |  |  |  |  |  |  |  |  |  |  |
| ≤19 | 15 | 32 | 44 | 60 | 73 | 96 | 116 | 120 | 121 | 116 | 104 |
| 20-39 | 30 | 58 | 82 | 87 | 109 | 133 | 155 | 183 | 174 | 174 | 152 |
| 40-59 | 14 | 35 | 53 | 62 | 70 | 69 | 81 | 79 | 92 | 83 | 80 |
| 60-69 | 2 | 4 | 9 | 13 | 18 | 19 | 19 | 23 | 22 | 21 | 13 |
| ≥70 | - | 1 | 4 | 9 | 11 | 11 | 16 | 20 | 23 | 15 | 9 |

**Supplementary Table 10.** Prevalent Testicular Cancer Cases Per Year Based on Filed Claims Within Period of Interest Alone (Without Lookback)

|  | 2011 | 2012 | 2013 | 2014 | 2015 | 2016 | 2017 | 2018 | 2019 | 2020 | 2021 |
| --- | --- | --- | --- | --- | --- | --- | --- | --- | --- | --- | --- |
|  | Frequency |  |  |  |  |  |  |  |  |  |  |
| Total | 63 | 78 | 75 | 100 | 123 | 124 | 169 | 176 | 146 | 142 | 141 |

|  | 2011 | 2012 | 2013 | 2014 | 2015 | 2016 | 2017 | 2018 | 2019 | 2020 | 2021 |
| --- | --- | --- | --- | --- | --- | --- | --- | --- | --- | --- | --- |
|  | Frequency |  |  |  |  |  |  |  |  |  |  |
| ≤19 | 15 | 20 | 16 | 31 | 32 | 37 | 57 | 47 | 43 | 51 | 48 |
| 20-39 | 31 | 33 | 30 | 35 | 52 | 59 | 61 | 80 | 58 | 56 | 65 |
| 40-59 | 14 | 22 | 20 | 23 | 30 | 18 | 35 | 29 | 33 | 28 | 25 |
| 60-69 | 3 | 2 | 6 | 6 | 6 | 7 | 6 | 12 | 6 | 5 | 2 |
| ≥70 | - | 1 | 3 | 5 | 3 | 3 | 10 | 8 | 6 | 2 | 1 |

**Supplementary Table 11.** Incident Testicular Cancer Cases Per Year Based on Paid Claims Only, Within Period of Interest.

|  | 2011 | 2012 | 2013 | 2014 | 2015 | 2016 | 2017 | 2018 | 2019 | 2020 | 2021 |
| --- | --- | --- | --- | --- | --- | --- | --- | --- | --- | --- | --- |
|  | Frequency |  |  |  |  |  |  |  |  |  |  |
| **Total** | **61** | **69** | **62** | **92** | **115** | **116** | **151** | **149** | **119** | **124** | **104** |
| Age, years |  |  |  |  |  |  |  |  |  |  |  |
| ≤19 | 15 | 17 | 12 | 28 | 29 | 37 | 48 | 35 | 31 | 41 | 29 |
| 20-39 | 30 | 28 | 24 | 31 | 49 | 52 | 53 | 71 | 45 | 53 | 49 |
| 40-59 | 14 | 21 | 18 | 22 | 28 | 17 | 34 | 26 | 31 | 23 | 23 |
| 60-69 | 2 | 2 | 5 | 6 | 6 | 7 | 6 | 10 | 6 | 5 | 2 |
| ≥70 | - | 1 | 3 | 5 | 3 | 3 | 10 | 7 | 6 | 2 | 1 |

**Supplementary Table 12.** Testicular Cancer Prevalence Across Years.

| Year | Total beneficiaries | Prevalence of filed claims (95% CI) | Prevalence of paid claims (95% CI) |
| --- | --- | --- | --- |
| 2011 | 14,702,032 | 0.00000429 (0.00000329–0.00000548) | 0.00000415 (0.00000317–0.00000533) |
| 2012 | 15,834,192 | 0.0000084 (0.00000703–0.00000995) | 0.00000821 (0.00000686–0.00000975) |
| 2013 | 16,945,352 | 0.0000116 (0.00001–0.0000133) | 0.0000113 (0.00000978–0.0000131) |
| 2014 | 44,403,501 | 0.00000529 (0.00000464–0.00000601) | 0.0000052 (0.00000455–0.00000592) |
| 2015 | 47,814,214 | 0.00000598 (0.00000531–0.00000672) | 0.00000588 (0.00000521–0.00000661) |
| 2016 | 47,960,271 | 0.00000692 (0.0000062–0.00000771) | 0.00000684 (0.00000612–0.00000762) |
| 2017 | 46,307,729 | 0.00000847 (0.00000765–0.00000935) | 0.00000836 (0.00000755–0.00000923) |
| 2018 | 51,486,361 | 0.00000851 (0.00000773–0.00000934) | 0.00000825 (0.00000749–0.00000908) |
| 2019 | 46,681,095 | 0.00000988 (0.00000899–0.0000108) | 0.00000925 (0.0000084–0.0000102) |
| 2020 | 46,024,677 | 0.00000956 (0.00000869–0.0000105) | 0.00000889 (0.00000805–0.00000979) |
| 2021 | 45,746,463 | 0.00000879 (0.00000795–0.00000969) | 0.00000783 (0.00000704–0.00000868) |

**Supplementary Table 13.** Prevalent Penile Cancer Cases Per Year Based on Paid Claims Only, Within Period of Interest and Lookback Period.

|  | 2011 | 2012 | 2013 | 2014 | 2015 | 2016 | 2017 | 2018 | 2019 | 2020 | 2021 |
| --- | --- | --- | --- | --- | --- | --- | --- | --- | --- | --- | --- |
|  | Frequency |  |  |  |  |  |  |  |  |  |  |
| **Total** | **11** | **26** | **44** | **58** | **75** | **82** | **89** | **84** | **97** | **96** | **76** |
| Age, years |  |  |  |  |  |  |  |  |  |  |  |
| ≤19 | - | - | - | - | - | - | - | - | - | - | - |
| 20-39 | 3 | 3 | 6 | 8 | 11 | 10 | 11 | 10 | 13 | 15 | 13 |
| 40-59 | 5 | 12 | 23 | 32 | 42 | 43 | 46 | 47 | 52 | 53 | 40 |
| 60-69 | 1 | 5 | 7 | 9 | 13 | 14 | 18 | 14 | 20 | 16 | 13 |
| ≥70 | 2 | 6 | 8 | 9 | 9 | 15 | 14 | 13 | 12 | 12 | 10 |

**Supplementary Table 14.**
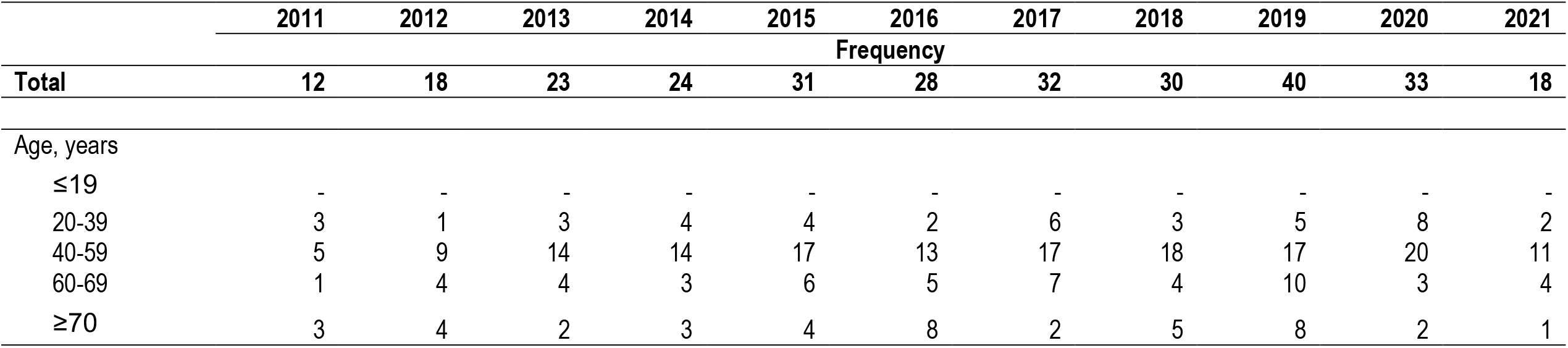
Prevalent Penile Cancer Cases Per Year Based on Filed Claims Within Period of Interest Alone (Without Lookback) 2011 2012 2013 2014 2015 2016 2017 2018 2019 2020 2021.

**Supplementary Table 15.** Incident Penile Cancer Cases Per Year Based on Paid Claims Only, Within Period of Interest.

|  | 2011 | 2012 | 2013 | 2014 | 2015 | 2016 | 2017 | 2018 | 2019 | 2020 | 2021 |
| --- | --- | --- | --- | --- | --- | --- | --- | --- | --- | --- | --- |
|  | Frequency |  |  |  |  |  |  |  |  |  |  |
| **Total** | **11** | **15** | **18** | **23** | **30** | **28** | **30** | **26** | **38** | **30** | **8** |
| Age, years |  |  |  |  |  |  |  |  |  |  |  |
| ≤19 | - | - | - | - | - | - | - | - | - | - | - |
| 20-39 | 3 | - | 3 | 4 | 4 | 2 | 5 | 3 | 4 | 8 | 1 |

|  | 2011 | 2012 | 2013 | 2014 | 2015 | 2016 | 2017 | 2018 | 2019 | 2020 | 2021 |
| --- | --- | --- | --- | --- | --- | --- | --- | --- | --- | --- | --- |
|  | Frequency |  |  |  |  |  |  |  |  |  |  |
| 40-59 | 5 | 7 | 11 | 13 | 16 | 13 | 16 | 18 | 17 | 18 | 5 |
| 60-69 | 1 | 4 | 2 | 3 | 6 | 5 | 7 | 2 | 10 | 2 | 1 |
| ≥70 | 2 | 4 | 2 | 3 | 4 | 8 | 2 | 3 | 7 | 2 | 1 |

**Supplementary Table 16.**
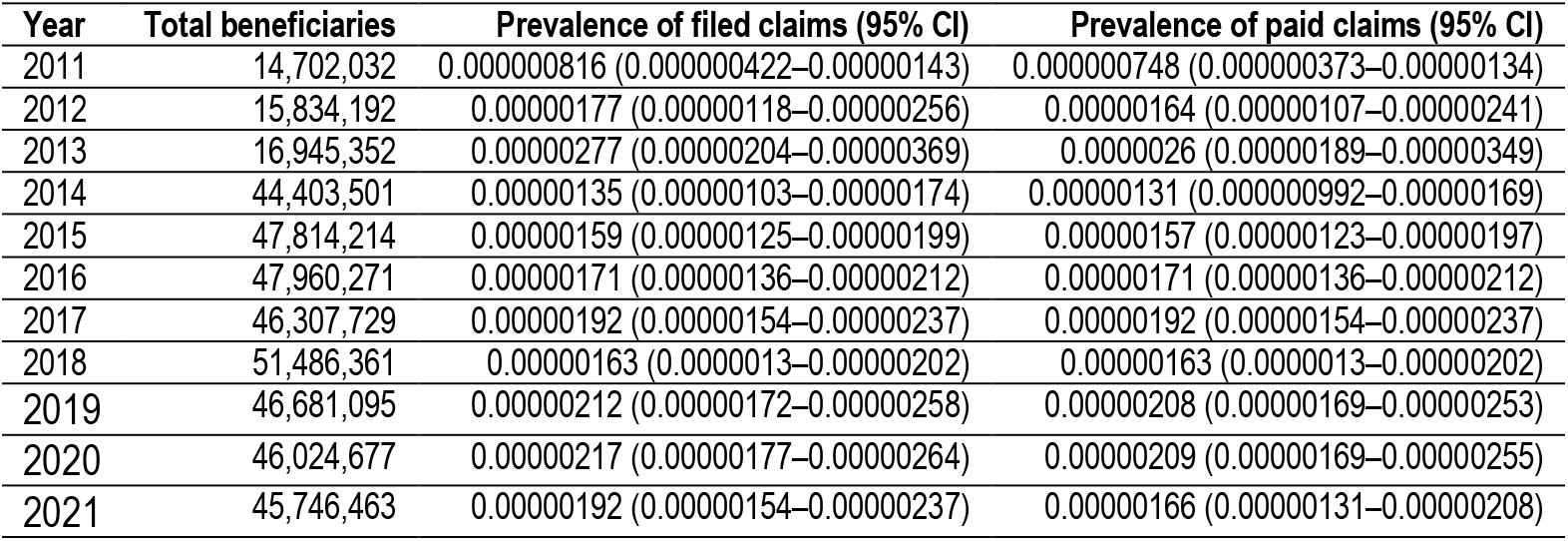
Penile Cancer Prevalence Across Years.

**Supplementary Table 17.** Prevalent Prostate Cancer Cases Per Year Based on Paid Claims Only Within Period of Interest and Lookback Period.

|  | 2011 | 2012 | 2013 | 2014 | 2015 | 2016 | 2017 | 2018 | 2019 | 2020 | 2021 |
| --- | --- | --- | --- | --- | --- | --- | --- | --- | --- | --- | --- |
|  | Frequency |  |  |  |  |  |  |  |  |  |  |
| **Total** | **835** | **1,657** | **2,446** | **2,611** | **2,965** | **3,348** | **3,620** | **3,765** | **3,771** | **3,486** | **2,844** |
| Age, years |  |  |  |  |  |  |  |  |  |  |  |
| ≤19 | 1 | 4 | 6 | 8 | 6 | 3 | 3 | 1 | 1 | 1 | 1 |
| 20-39 | 6 | 11 | 14 | 9 | 7 | 3 | 6 | 5 | 6 | 4 | 5 |
| 40-59 | 74 | 144 | 221 | 241 | 255 | 272 | 280 | 310 | 308 | 314 | 261 |
| 60-69 | 238 | 473 | 729 | 770 | 881 | 964 | 1,072 | 1,113 | 1,133 | 1,051 | 882 |
| ≥70 | 516 | 1,025 | 1,476 | 1,583 | 1,816 | 2,106 | 2,259 | 2,336 | 2,323 | 2,116 | 1,695 |

|  | 2011 | 2012 | 2013 | 2014 | 2015 | 2016 | 2017 | 2018 | 2019 | 2020 | 2021 |
| --- | --- | --- | --- | --- | --- | --- | --- | --- | --- | --- | --- |
|  | Frequency |  |  |  |  |  |  |  |  |  |  |
| No | 793 | 1,496 | 2,168 | 2,384 | 2,837 | 3,339 | 3,615 | 3,752 | 3,754 | 3,466 | 2,832 |
| Yes | 42 | 161 | 278 | 227 | 128 | 9 | 5 | 13 | 17 | 20 | 12 |
| **Region** |  |  |  |  |  |  |  |  |  |  |  |
| NCR | 236 | 493 | 745 | 790 | 818 | 842 | 865 | 915 | 898 | 765 | 556 |
| I – Ilocos | 58 | 110 | 174 | 195 | 236 | 260 | 282 | 283 | 322 | 322 | 291 |
| II – Cagayan | 35 | 56 | 86 | 83 | 110 | 126 | 145 | 149 | 147 | 157 | 134 |
| III – Central Luzon | 73 | 150 | 223 | 232 | 251 | 276 | 317 | 341 | 343 | 321 | 246 |
| IV-A - CALABARZON | 46 | 85 | 122 | 128 | 155 | 184 | 202 | 205 | 217 | 191 | 170 |
| IV-B – MIMAROPA | 37 | 72 | 111 | 119 | 140 | 156 | 169 | 178 | 177 | 170 | 143 |
| V – BICOL | 27 | 47 | 68 | 70 | 96 | 123 | 133 | 137 | 140 | 153 | 140 |
| VI – Western Visayas | 79 | 149 | 191 | 197 | 228 | 276 | 301 | 308 | 319 | 315 | 281 |
| VII – Central Visayas | 106 | 207 | 297 | 279 | 288 | 318 | 342 | 366 | 341 | 323 | 261 |
| VIII – Eastern Visayas | 7 | 21 | 32 | 47 | 68 | 91 | 107 | 121 | 129 | 124 | 93 |
| IX – Zamboanga Peninsula | 13 | 29 | 36 | 57 | 65 | 86 | 91 | 90 | 91 | 73 | 60 |
| X – Northern Mindanao | 27 | 50 | 80 | 102 | 145 | 183 | 193 | 191 | 192 | 177 | 142 |
| XI – Davao Region | 48 | 86 | 126 | 140 | 164 | 186 | 173 | 180 | 167 | 147 | 104 |
| XII - Soccsksargen | 18 | 43 | 67 | 69 | 90 | 108 | 143 | 153 | 155 | 136 | 120 |
| CAR | 16 | 42 | 65 | 74 | 74 | 85 | 108 | 107 | 94 | 73 | 64 |
| ARMM | - | 1 | 2 | 4 | 4 | 3 | 2 | 1 | 2 | 3 | 3 |
| CARAGA | 8 | 13 | 18 | 21 | 32 | 43 | 45 | 37 | 35 | 34 | 36 |
| UNKNOWN | 1 | 3 | 3 | 4 | 1 | 2 | 2 | 3 | 2 | 2 | - |
| **Class** |  |  |  |  |  |  |  |  |  |  |  |
| Ambulatory surgical clinic | - | - | 9 | 8 | 8 | 1 | - | - | - | - | - |
| Infirmery / dispensary | 10 | 23 | 41 | 34 | 23 | 13 | 24 | 28 | 24 | 14 | 8 |
| Level 1 | 124 | 233 | 359 | 402 | 494 | 574 | 641 | 645 | 655 | 659 | 604 |
| Level 2 | 269 | 499 | 722 | 803 | 1,021 | 1,239 | 1,355 | 1,402 | 1,407 | 1,323 | 1,126 |

|  | 2011 | 2012 | 2013 | 2014 | 2015 | 2016 | 2017 | 2018 | 2019 | 2020 | 2021 |
| --- | --- | --- | --- | --- | --- | --- | --- | --- | --- | --- | --- |
|  | Frequency |  |  |  |  |  |  |  |  |  |  |
| Level 3 | 431 | 899 | 1,312 | 1,360 | 1,418 | 1,519 | 1,598 | 1,687 | 1,683 | 1,488 | 1,106 |
| Unknown | 1 | 3 | 3 | 4 | 1 | 2 | 2 | 3 | 2 | 2 | - |

**Supplementary Table 18.**
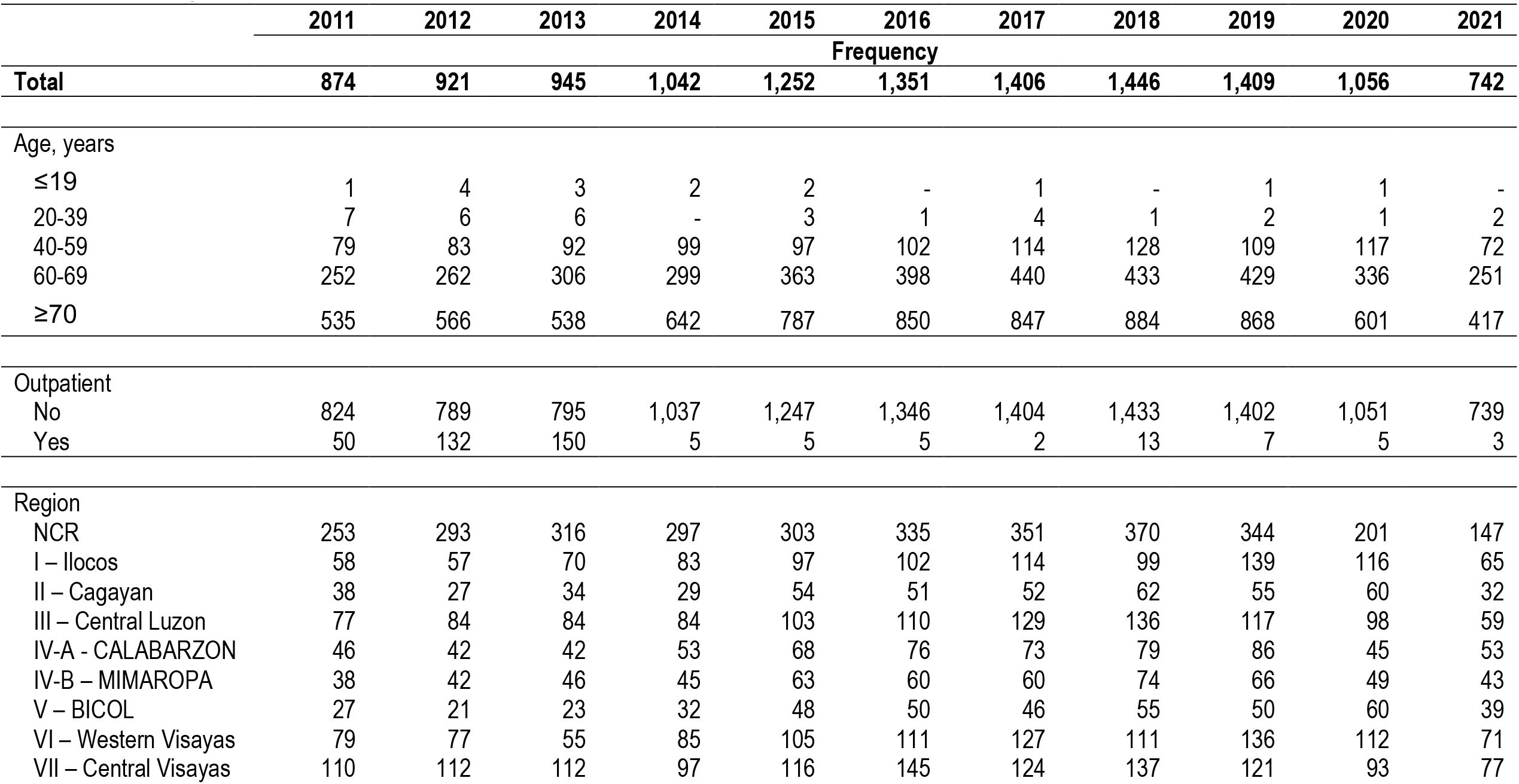
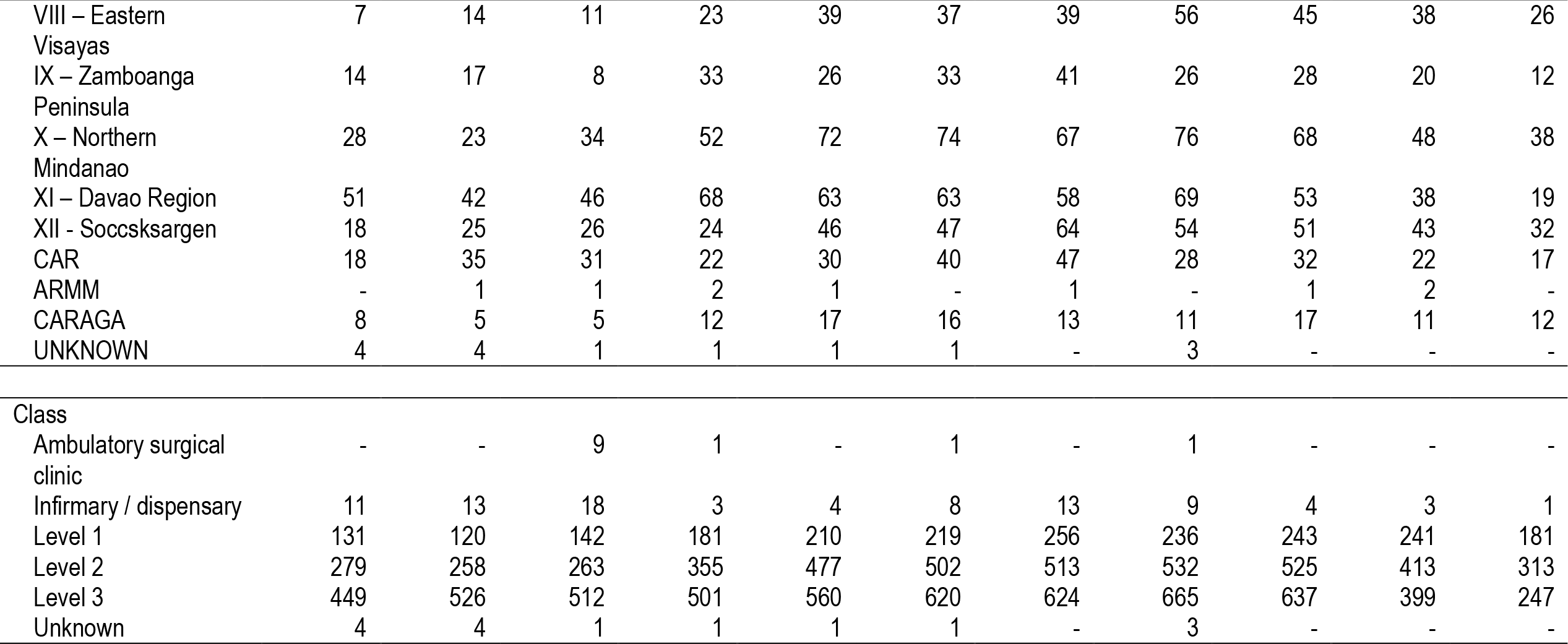
Prevalent Prostate Cancer Cases Per Year Based on Filed Claims Within Period of Interest Along (Without Lookback Period)

|  | 2011 | 2012 | 2013 | 2014 | 2015 | 2016 | 2017 | 2018 | 2019 | 2020 | 2021 |
| --- | --- | --- | --- | --- | --- | --- | --- | --- | --- | --- | --- |
|  | Frequency |  |  |  |  |  |  |  |  |  |  |
| **Total** | **874** | **921** | **945** | **1,042** | **1,252** | **1,351** | **1,406** | **1,446** | **1,409** | **1,056** | **742** |
| Age, years |  |  |  |  |  |  |  |  |  |  |  |
| ≤19 | 1 | 4 | 3 | 2 | 2 | - | 1 | - | 1 | 1 | - |
| 20-39 | 7 | 6 | 6 | - | 3 | 1 | 4 | 1 | 2 | 1 | 2 |
| 40-59 | 79 | 83 | 92 | 99 | 97 | 102 | 114 | 128 | 109 | 117 | 72 |
| 60-69 | 252 | 262 | 306 | 299 | 363 | 398 | 440 | 433 | 429 | 336 | 251 |
| ≥70 | 535 | 566 | 538 | 642 | 787 | 850 | 847 | 884 | 868 | 601 | 417 |
| Outpatient |  |  |  |  |  |  |  |  |  |  |  |
| No | 824 | 789 | 795 | 1,037 | 1,247 | 1,346 | 1,404 | 1,433 | 1,402 | 1,051 | 739 |
| Yes | 50 | 132 | 150 | 5 | 5 | 5 | 2 | 13 | 7 | 5 | 3 |
| Region |  |  |  |  |  |  |  |  |  |  |  |
| NCR | 253 | 293 | 316 | 297 | 303 | 335 | 351 | 370 | 344 | 201 | 147 |
| I – Ilocos | 58 | 57 | 70 | 83 | 97 | 102 | 114 | 99 | 139 | 116 | 65 |
| II – Cagayan | 38 | 27 | 34 | 29 | 54 | 51 | 52 | 62 | 55 | 60 | 32 |
| III – Central Luzon | 77 | 84 | 84 | 84 | 103 | 110 | 129 | 136 | 117 | 98 | 59 |
| IV-A - CALABARZON | 46 | 42 | 42 | 53 | 68 | 76 | 73 | 79 | 86 | 45 | 53 |
| IV-B – MIMAROPA | 38 | 42 | 46 | 45 | 63 | 60 | 60 | 74 | 66 | 49 | 43 |
| V – BICOL | 27 | 21 | 23 | 32 | 48 | 50 | 46 | 55 | 50 | 60 | 39 |
| VI – Western Visayas | 79 | 77 | 55 | 85 | 105 | 111 | 127 | 111 | 136 | 112 | 71 |
| VII – Central Visayas | 110 | 112 | 112 | 97 | 116 | 145 | 124 | 137 | 121 | 93 | 77 |

|  | 2011 | 2012 | 2013 | 2014 | 2015 | 2016 | 2017 | 2018 | 2019 | 2020 | 2021 |
| --- | --- | --- | --- | --- | --- | --- | --- | --- | --- | --- | --- |
|  | Frequency |  |  |  |  |  |  |  |  |  |  |
| VIII – Eastern Visayas | 7 | 14 | 11 | 23 | 39 | 37 | 39 | 56 | 45 | 38 | 26 |
| IX – Zamboanga Peninsula | 14 | 17 | 8 | 33 | 26 | 33 | 41 | 26 | 28 | 20 | 12 |
| X – Northern Mindanao | 28 | 23 | 34 | 52 | 72 | 74 | 67 | 76 | 68 | 48 | 38 |
| XI – Davao Region | 51 | 42 | 46 | 68 | 63 | 63 | 58 | 69 | 53 | 38 | 19 |
| XII - Soccsksargen | 18 | 25 | 26 | 24 | 46 | 47 | 64 | 54 | 51 | 43 | 32 |
| CAR | 18 | 35 | 31 | 22 | 30 | 40 | 47 | 28 | 32 | 22 | 17 |
| ARMM | - | 1 | 1 | 2 | 1 | - | 1 | - | 1 | 2 | - |
| CARAGA | 8 | 5 | 5 | 12 | 17 | 16 | 13 | 11 | 17 | 11 | 12 |
| UNKNOWN | 4 | 4 | 1 | 1 | 1 | 1 | - | 3 | - | - | - |
| **Class** |  |  |  |  |  |  |  |  |  |  |  |
| Ambulatory surgical clinic | - | - | 9 | 1 | - | 1 | - | 1 | - | - | - |
| Infirmery / dispensary | 11 | 13 | 18 | 3 | 4 | 8 | 13 | 9 | 4 | 3 | 1 |
| Level 1 | 131 | 120 | 142 | 181 | 210 | 219 | 256 | 236 | 243 | 241 | 181 |
| Level 2 | 279 | 258 | 263 | 355 | 477 | 502 | 513 | 532 | 525 | 413 | 313 |
| Level 3 | 449 | 526 | 512 | 501 | 560 | 620 | 624 | 665 | 637 | 399 | 247 |
| Unknown | 4 | 4 | 1 | 1 | 1 | 1 | - | 3 | - | - | - |

**Supplementary Table 19.** Incident Prostate Cancer Cases Per Year Based on Paid Claims Only, Within Period of Interest and Lookback Period.

|  | 2011 | 2012 | 2013 | 2014 | 2015 | 2016 | 2017 | 2018 | 2019 | 2020 | 2021 |
| --- | --- | --- | --- | --- | --- | --- | --- | --- | --- | --- | --- |
|  | Frequency |  |  |  |  |  |  |  |  |  |  |
| **Total** | **835** | **822** | **789** | **919** | **1,132** | **1,189** | **1,203** | **1,216** | **1,168** | **918** | **605** |
| **Age, years** |  |  |  |  |  |  |  |  |  |  |  |
| ≤19 | 1 | 3 | 2 | 2 | 2 | 0 | 1 | 0 | 0 | 1 | 0 |
| 20-39 | 6 | 3 | 5 | 0 | 2 | 0 | 3 | 1 | 2 | 1 | 2 |

|  | 2011 | 2012 | 2013 | 2014 | 2015 | 2016 | 2017 | 2018 | 2019 | 2020 | 2021 |
| --- | --- | --- | --- | --- | --- | --- | --- | --- | --- | --- | --- |
|  | Frequency |  |  |  |  |  |  |  |  |  |  |
| 40-59 | 74 | 71 | 76 | 88 | 84 | 93 | 100 | 115 | 89 | 106 | 57 |
| 60-69 | 238 | 237 | 260 | 258 | 326 | 346 | 374 | 350 | 352 | 289 | 199 |
| ≥70 | 516 | 508 | 446 | 571 | 718 | 750 | 725 | 750 | 725 | 521 | 347 |
| Outpatient |  |  |  |  |  |  |  |  |  |  |  |
| No | 793 | 706 | 674 | 916 | 1,130 | 1,188 | 1,202 | 1,207 | 1,163 | 915 | 603 |
| Yes | 42 | 116 | 115 | 3 | 2 | 1 | 1 | 9 | 5 | 3 | 2 |
| Region |  |  |  |  |  |  |  |  |  |  |  |
| NCR | 236 | 254 | 254 | 266 | 258 | 288 | 286 | 302 | 262 | 157 | 107 |
| I – Ilocos | 58 | 54 | 64 | 73 | 90 | 89 | 98 | 86 | 124 | 102 | 53 |
| II – Cagayan | 35 | 22 | 29 | 23 | 50 | 48 | 46 | 51 | 42 | 52 | 29 |
| III – Central Luzon | 73 | 77 | 73 | 74 | 96 | 101 | 115 | 117 | 99 | 89 | 46 |
| IV-A - CALABARZON | 46 | 39 | 35 | 49 | 65 | 66 | 68 | 67 | 75 | 38 | 48 |
| IV-B – MIMAROPA | 37 | 35 | 36 | 38 | 57 | 54 | 53 | 59 | 55 | 43 | 38 |
| V – BICOL | 27 | 20 | 21 | 28 | 46 | 46 | 40 | 44 | 46 | 55 | 34 |
| VI – Western Visayas | 79 | 70 | 44 | 77 | 94 | 96 | 103 | 91 | 107 | 99 | 59 |
| VII – Central Visayas | 106 | 101 | 92 | 77 | 103 | 123 | 102 | 120 | 100 | 85 | 59 |
| VIII – Eastern Visayas | 7 | 13 | 11 | 22 | 35 | 33 | 35 | 48 | 41 | 28 | 19 |
| IX – Zamboanga Peninsula | 13 | 16 | 6 | 33 | 24 | 28 | 35 | 21 | 27 | 19 | 12 |
| X – Northern Mindanao | 27 | 23 | 29 | 47 | 65 | 66 | 59 | 64 | 62 | 45 | 31 |
| XI – Davao Region | 48 | 39 | 40 | 60 | 61 | 60 | 51 | 64 | 44 | 35 | 16 |
| XII - Soccsksargen | 18 | 25 | 25 | 18 | 43 | 39 | 59 | 47 | 44 | 39 | 30 |
| CAR | 16 | 26 | 24 | 20 | 27 | 36 | 41 | 24 | 25 | 20 | 13 |
| ARMM | - | 1 | 1 | 2 | 1 | - | 1 | - | 1 | 2 | - |
| CARAGA | 8 | 5 | 5 | 11 | 16 | 15 | 11 | 9 | 14 | 10 | 11 |
| UNKNOWN | 1 | 2 | - | 1 | 1 | 1 | - | 2 | - | - | - |

|  | 2011 | 2012 | 2013 | 2014 | 2015 | 2016 | 2017 | 2018 | 2019 | 2020 | 2021 |
| --- | --- | --- | --- | --- | --- | --- | --- | --- | --- | --- | --- |
|  | Frequency |  |  |  |  |  |  |  |  |  |  |
| Ambulatory surgical clinic | - | - | 9 | 1 | - | - | - | - | - | - | - |
| Infirmar y / dispensary | 10 | 11 | 17 | 2 | 3 | 7 | 12 | 7 | 4 | 3 | 1 |
| Level 1 | 124 | 108 | 127 | 153 | 197 | 197 | 223 | 197 | 204 | 218 | 148 |
| Level 2 | 269 | 235 | 219 | 321 | 437 | 439 | 444 | 448 | 442 | 363 | 265 |
| Level 3 | 431 | 466 | 417 | 441 | 494 | 545 | 524 | 562 | 518 | 334 | 191 |
| Unknown | 1 | 2 | - | 1 | 1 | 1 | - | 2 | - | - | - |

**Supplementary Table 20.** Prostate Cancer Prevalence Across Years.

| Year | Total beneficiaries | Prevalence of filed claims (95% CI) | Prevalence of paid claims (95% CI) |
| --- | --- | --- | --- |
| 2011 | 14,702,032 | 0.0000594 (0.0000556–0.0000635) | 0.0000568 (0.000053–0.0000608) |
| 2012 | 15,834,192 | 0.0001084 (0.0001034–0.0001137) | 0.0001046 (0.0000997–0.0001098) |
| 2013 | 16,945,352 | 0.0001493 (0.0001435–0.0001552) | 0.0001443 (0.0001387–0.0001502) |
| 2014 | 44,403,501 | 0.0000603 (0.000058–0.0000626) | 0.0000588 (0.0000566–0.0000611) |
| 2015 | 47,814,214 | 0.0000634 (0.0000612–0.0000657) | 0.000062 (0.0000598–0.0000643) |
| 2016 | 47,960,271 | 0.0000709 (0.0000685–0.0000733) | 0.0000698 (0.0000675–0.0000722) |
| 2017 | 46,307,729 | 0.0000796 (0.000077–0.0000822) | 0.0000782 (0.0000756–0.0000808) |
| 2018 | 51,486,361 | 0.0000749 (0.0000725–0.0000773) | 0.0000731 (0.0000708–0.0000755) |
| 2019 | 46,681,095 | 0.0000841 (0.0000815–0.0000868) | 0.0000808 (0.0000782–0.0000834) |
| 2020 | 46,024,677 | 0.0000792 (0.0000767–0.0000818) | 0.0000757 (0.0000732–0.0000783) |
| 2021 | 45,746,463 | 0.0000659 (0.0000636–0.0000683) | 0.0000622 (0.0000599–0.0000645) |

